# A Personalized Whole-Food Diet Differentially Modulates Glucoregulatory and Cognitive Responses Compared With Conventional Dietary Counseling in Young Black and White Adults With Overweight or Obesity: An 8-Week Randomized Controlled Trial

**DOI:** 10.64898/2026.05.27.26354244

**Authors:** Onyedika G. Ani, Elaheh Rabbani, Jaapna Dhillon

**Affiliations:** Division of Food, Nutrition, and Exercise Sciences, College of Agriculture, Food and Natural Resources (CAFNR), University of Missouri-Columbia; Department of Nutrition and Exercise Physiology, School of Medicine, University of Missouri-Columbia, MO

**Author notes:** Corresponding Author: Jaapna Dhillon, PhD, Department of Nutrition and Exercise Physiology School of Medicine, University of Missouri, Columbia 312 Gwynn Hall, Columbia, MO 65211.

**Keywords:** dietary intervention, personalized nutrition, diet quality, oral glucose tolerance test, insulin resistance, racial differences, obesity, attention

## Abstract

**Background:** Black adults bear a disproportionate burden of cardiometabolic dysfunction, yet most dietary trial evidence comes from predominantly White cohorts.

**Objective:** To evaluate whether a personalized whole-food dietary intervention improves cardiometabolic outcomes more in Black than White young adults with overweight or obesity.

**Methods:** In this 8-week randomized, controlled trial (ClinicalTrials.gov: NCT04635917), 112 Black and White adults (18-35 years; BMI 25-45 kg/m²) were block-randomized by race to a personalized dietary intervention providing whole foods (PD, n=57) or conventional dietary counseling at baseline (BL) using MyPlate guidelines (CD, n=55). Primary outcomes were Matsuda Index and fasting and OGTT-derived glucose, insulin, and non-esterified fatty acids. Other glucoregulatory, cardiovascular, anthropometric, appetite, and cognitive outcomes were also assessed. Outcomes were analyzed using baseline-adjusted linear models with sensitivity analyses adjusting for baseline BMI and food security score.

**Results:** Compliance with study food consumption was 85-91%. Diet quality was higher in PD than CD (P < 0.05), with larger gains in vegetable-related outcomes among Black participants (group x race, P < 0.05). HOMA-β was lower in PD than CD overall (P < 0.05). In sensitivity analyses, Black PD participants had greater fasting insulin reductions than White, especially in the latter half of intervention (week x group x race, P < 0.05), with a similar tendency for HOMA-IR. Glucose AUC 0-30 min was higher in White than Black PD participants (group x race, P < 0.05). Concentration performance was higher in PD than CD overall (P < 0.05), with larger gains in processing speed and accuracy among Black than White participants (group x race, P < 0.05). No effects were observed for cardiovascular or appetite outcomes.

**Conclusions:** The personalized whole-food intervention produced differential effects in fasting insulin and early-phase glucose handling, and greater benefits in attention, in Black compared with White young adults with overweight or obesity during weight maintenance.

**Clinical Trial Registry number:** ClinicalTrials.gov ID: NCT04635917

## INTRODUCTION

Black adults in the U.S. exhibit a disproportionate burden of cardiometabolic dysfunction, including higher prevalence of obesity (1.3-fold), hypertension (1.3-fold), and diabetes (1.6-fold), compared with non-Hispanic White adults (1,2). Racial disparities in cardiometabolic health have persisted over time, with increases in optimal cardiometabolic health observed among non-Hispanic White adults in recent decades but decreases among non-Hispanic Black adults (3). Race-differential glucoregulatory features have been documented across the glycemic spectrum, including lower insulin sensitivity and insulin clearance alongside higher insulin secretion and disposition index in normoglycemic Black compared with non-Hispanic White adults at familial risk for diabetes (4), and a higher prevalence of less favorable OGTT glucose curve shapes among Black compared with non-Hispanic White adults with early type 2 diabetes (5). Although overweight and obesity are major contributors to cardiometabolic risk, racial differences in cardiometabolic risk are not fully explained by BMI-defined body weight alone (6).

Nearly half of cardiometabolic deaths in the U.S. are associated with suboptimal diets (7). Higher diet quality is commonly reflected in adherence to healthy dietary patterns and diet quality indices such as the Mediterranean (MED) diet, Dietary Approaches to Stop Hypertension (DASH), Healthy Eating Index (HEI), and Alternative Healthy Eating Index (AHEI), which generally emphasize fruits, vegetables, legumes, nuts, whole grains, and minimally processed foods while limiting saturated fat, sodium, and added sugar. In meta-analyses of observational studies, higher diet quality is consistently associated with more favorable cardiometabolic profiles and lower risk of cardiovascular disease, type 2 diabetes, and metabolic syndrome (8–17).

However, the cohorts underlying this dietary evidence base remain predominantly White, and race-stratified evidence from randomized controlled trials is limited. A systematic review showed that only 34 dietary intervention trials reported analyses stratified by race or ethnicity (18). Observed differences were most consistent for blood pressure with several DASH-style and sodium-reduction trials suggesting greater blood pressure responsiveness among Black compared with White adults (19–23). Lipid responses were largely comparable, while triglyceride and apolipoprotein findings were less consistent. Evidence for glucoregulatory outcomes was limited and mixed, including lower insulin clearance among Black participants in one study (24), a diet-specific difference in beta-cell response in another (25), and similar reductions in insulin resistance between Black and White participants in a low-fat vegan diet trial, though in the context of weight loss (26).

Interpretation of this mixed evidence base is further complicated by the differential burden of food insecurity in Black (22.4%) compared with White households (9.3%) (27). Food insecurity is independently associated with lower diet quality and greater risk of type 2 diabetes, hypertension, and incident cardiovascular disease (28–30). Hence, in free-living populations, race differences in cardiometabolic outcomes may therefore reflect differences in food access, race-related genetic or metabolic factors that interact with diet, or both, and these can only be disentangled when dietary exposure is standardized across groups, as is achieved through direct food provision in a controlled trial.

Accordingly, the present 8-week randomized, controlled, parallel-arm trial was conducted in Black and non-Hispanic White adults with overweight or obesity aged 18 to 35 years. This age range represents a period during which early metabolic dysfunction may already be detectable, but before cardiometabolic disease is typically established, providing a potential window for dietary intervention to influence long-term cardiometabolic risk trajectories (31,32). This study evaluated whether improved diet quality, delivered through provision of a personalized whole-food intervention (PD), improves cardiometabolic outcomes, with glucoregulatory outcomes as primary, compared with conventional dietary advice (CD). Cardiovascular, appetite, and cognitive outcomes were also evaluated, with cognitive outcomes included given that insulin resistance has been shown to predict cognitive decline in adults (33,34). We hypothesized that glucoregulatory outcomes and related secondary cardiovascular, appetite, and cognitive outcomes would be more favorable in PD than CD over the intervention period, with more favorable effects of PD in Black than non-Hispanic White participants.

## METHODS

### Study Approval

The human trial procedures and data management of this study were approved by the University of Missouri Institutional Review Board (IRB No. 2022561). The study was registered on ClinicalTrials.gov (NCT04635917) on November 2020. There was no patient or public involvement in any aspects of the trial and no major changes were made to the trial protocol after commencement.

### Study Design

This study was an 8-week randomized, controlled, parallel-arm clinical trial conducted at the University of Missouri-Columbia. Participants were block-randomized by race (Black or White) into one of two intervention arms in a 1:1 ratio: (1) Personalized Whole-Food Diet (PD) or (2) Conventional Dietary Advice (CD). This study used a single-blind design in which specific laboratory personnel were unaware of participants’ group assignments. Sample blinding was maintained by labeling specimens with coded identification numbers. The study investigator generated the allocation sequence using computer-based block randomization stratified by race. Within each stratum, allocation lists were randomly ordered using the RAND() function in Microsoft Excel, and eligible participants were assigned to study groups in sequence following enrollment confirmation.

Participants (n = 57) in the PD group received individualized nutrition counseling from a registered dietitian focused on three goals: (a) increasing fiber intake to meet minimum recommendations, (b) increasing unsaturated-to-saturated fat ratio to >2.5, and (c) a personalized goal based on each participant’s baseline diet and metabolic phenotype (including body weight, fasting glucose, HbA1c, and lipid profile). The personalized goal most commonly focused on reducing added sugar intake (particularly from sugar-sweetened beverages, soda, juice, and sweets) and limiting processed foods, often alongside complementary goals such as increasing intake of whole foods, improving meal patterns (e.g., eating breakfast, avoiding skipped meals), and supporting adherence to study foods. PD participants attended virtual counseling sessions (30 to 60 minutes) at weeks 1, 2, 3, 4, 6, and 8. PD group was provided the following foods for 8 weeks to help with adherence, walnuts, almonds, prunes, clementines, carrots, broccoli, and sweet peppers, consumed daily. Daily quantities and their nutritional composition are outlined in **Supplemental Table 1.**

Participants (n = 55) in the CD group received non-personalized, conventional dietary advice from the same registered dietitian based on the MyPlate guidelines (35) at the start of the study only (week 1). To maintain the same contact frequency with study staff as the PD group, CD participants met with a dietitian at weeks 2, 3, 4, 6, and 8 for 24-hour dietary recalls without additional counseling.

### Sample Size Calculation

The sample size for the present study was informed by effect sizes observed in a prior 8-week almond consumption study from our group, in which the Matsuda Index differed between the almond intervention and cracker control groups (pooled mean ± SD: 7.8 ± 5.2 vs. 4.3 ± 2.5; 82% between-group difference; Cohen’s d = 0.85; n = 20 per group; p < 0.05) (36). With approximately 23 participants per race x group cell and approximately 46 per main effect group in the present study, at α = 0.05 (two-sided) and power = 0.80, the minimum detectable Cohen’s d was approximately 0.58 for main effects of Group (PD vs. CD) or Race (Black vs. White) and approximately 0.83 for cell-level pairwise comparisons and Group x Race interaction contrasts.

### Participants

#### Recruitment, Inclusion, and Exclusion Criteria

Participants were recruited between December 22, 2020 and December 21, 2024 through public advertisements and completed pre-screening forms to determine eligibility. Inclusion criteria were: (a) male and female adults aged 18 to 35 years; (b) self-identified race of Black or non-Hispanic White; (c) BMI of 25 to 45 kg/m²; (d) willingness to consume study foods and comply with the study protocol; (e) consistent diet and activity patterns for at least four weeks prior to enrollment; (f) weight-stable (≤5 kg change over the prior 3 months); and (g) non-smoker for >1 year. Exclusion criteria were: (a) allergies to fruits, vegetables, or nuts provided in the study; (b) illicit drug use; (c) recent initiation of medications affecting metabolism or appetite; (d) recent antibiotic use; (e) uncontrolled hypertension (blood pressure ≥180/110 mmHg); (f) diabetes; (g) drug therapy for coronary artery disease, peripheral artery disease, congestive heart failure, or dyslipidemia; (h) gastrointestinal disease and/or history of bariatric surgery; (i) HIV positivity; and (j) pregnancy or lactation.

Of the 154 individuals screened for eligibility, 112 were randomized, and 93 completed all study visits. The detailed participant CONSORT flow is shown in **Figure 1**.

**Figure 1.**
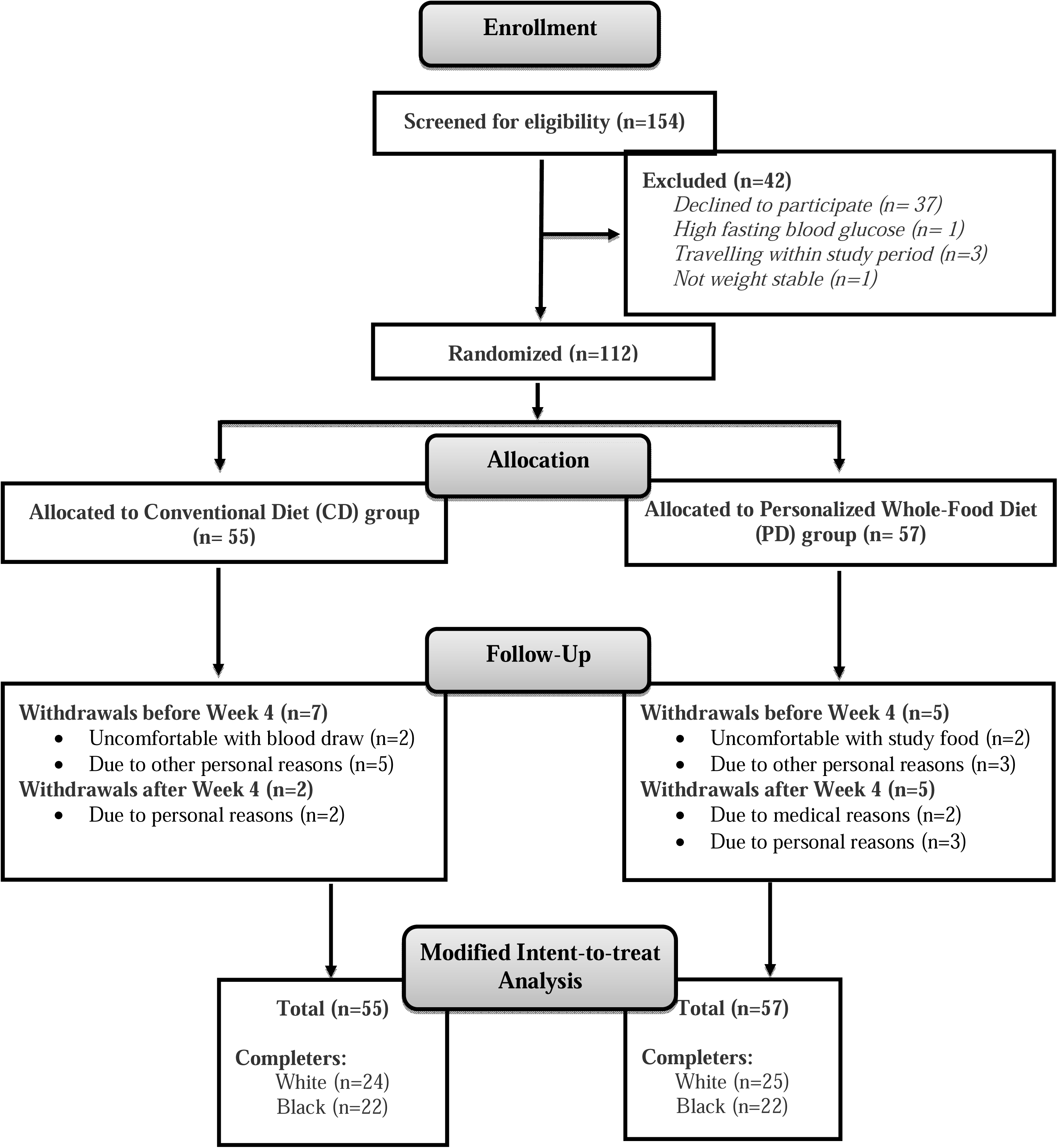
CONSORT flow diagram of Black and White participants in the PD and CD groups from enrollment to 8-week study completion.

### Screening Measures

Prior to randomization, participants completed assessment of baseline metabolic phenotype to confirm inclusion criteria and to inform personalization of the third dietary goal for participants assigned to the PD group. Phenotyping included body weight, fasting glucose by point-of-care glucometer, HbA1c by DCA Vantage Analyzer (Siemens, UK) or Quest Diagnostics laboratory analysis, and fasting lipid profile by Cholestech LDX (Abbott, USA). No interim analyses or stopping guidelines were planned.

### Baseline Participant Characteristics

Household food security was assessed at baseline using the U.S. Household Food Security Survey Module: Six-Item Short Form developed by the U.S. Department of Agriculture, Economic Research Service (37). Scores were calculated according to standard USDA scoring guidelines and were used as a covariate in pre-specified sensitivity analyses.

### Study Visits and Schedule

The 8-week study involved three data collection points (baseline (BL), week 4 (W4), and week 8 (W8)), three check-in visits (W2, W3, W6). One to two weeks before the start of the intervention, participants entered a phase-in period during which they maintained a consistent diet and physical activity routine and were fully informed of all study procedures.

At BL, W4, and W8 visits, participants arrived at the study clinic in the morning after an 8 to 12 hour fast. They were seated and rested for approximately five minutes before measurements began. Blood samples were collected at BL, W4, and W8 into serum separator tubes (SST). Whole blood was held at room temperature for approximately 1 hour, then centrifuged for 10 minutes at 1.3 RCF. Serum was transferred into labeled microcentrifuge tubes and stored at −80°C until analysis.

### Intervention Adherence and Acceptance

#### Food Consumption

Participants reported their daily food consumption by responding to text messages sent on weekdays. Weekly food supplies were provided in daily portions and participants were encouraged to consume each portion daily (at least 5 times per week). For each participant, food consumption data were organized by food type and day, with each food coded as consumed or not consumed on each reporting day.

### Dietary Goal Attainment (PD group only)

During the initial counseling session, the three dietary goals were established for each PD participant and documented in the dietitian’s notes. The dietitian used the ASA24® Dietary Assessment Tool to collect 24-hour dietary recalls (38). Goal attainment was assessed during follow-up visits at weeks 2, 3, 4, 6, and 8. For each participant, goal attainment data were organized by goal and visit, with each goal coded at each visit as achieved or not achieved.

### Counseling Visit Attendance

Attendance at scheduled counseling and recall visits was tracked for participants in both groups across the six time points (W1, W2, W3, W4, W6, W8). At each time point, attendance was recorded as attended or not attended based on the presence of recorded visit data.

### Food Acceptance and Palatability

Biweekly acceptance of each study food was evaluated using a 9-point food action (FACT) rating scale (1 = “I would eat this if I were forced to”; 9 = “I would eat this at every opportunity”) (39). Palatability was assessed biweekly using a hedonic general labeled magnitude scale (gLMS), with 0 = “extremely unpalatable” and 100 = “extremely palatable” (40).

### Lifestyle Behaviors and Appetite

#### Dietary Intake

Habitual dietary intake was assessed using the National Cancer Institute’s semi-quantitative Diet History Questionnaire III (DHQ III), a one-month food frequency questionnaire, administered at BL, W4, and W8. Total and detailed nutrient compositions, food group intakes, and diet quality scores were derived from DHQ III outputs. Healthy Eating Index (HEI-2015), Alternative Healthy Eating Index (AHEI), Dietary Approaches to Stop Hypertension (DASH), and Mediterranean (MED) diet pattern indices were calculated using the dietaryindex package in R (42).

### Physical Activity

Physical activity was assessed at BL, W4, and W8 using a waist-worn ActiGraph® accelerometer. Participants were instructed to wear the device on one weekday and one weekend day at each assessment period. Accelerometer data were processed using ActiLife 6® Data Scoring Grid software with a 10-second epoch length. Non-wear time was identified using the Troiano (2007) algorithm (43). Energy expenditure and metabolic equivalent (MET) values were estimated using the Freedson Combination (1998) (44), and Freedson Adult algorithms (default in ActiLife software), with time spent in sedentary, light, moderate, and vigorous activity classified using the Freedson (1998) MET-based thresholds. Average physical activity values were calculated as the mean of weekday and weekend measurements.

### Sleep

Sleep duration was assessed by daily participant self-report via text message. Each day, participants reported the total number of hours they slept the previous night. Sleep duration was summarized by averaging daily reports from W1, which served as the baseline sleep measure, W4/W5, which served as the midpoint sleep measure, and W8, which served as the end-of-intervention sleep measure.

### Free-Living Appetite Ratings

Participants completed hourly appetite ratings via text-message prompts from 8:00 am to 11:00 pm at BL and W8. The questionnaire assessed hunger, fullness, desire to eat, and prospective consumption using 100-mm visual analog scales ranging from “not at all” (0) to “extremely” (100) (45). Time points with high rates of missingness (e.g., 11:00 pm) were excluded from analysis prior to imputation, as imputation under these conditions would not yield meaningful values. The AUC for morning, afternoon, evening, and overall 14-hour periods was calculated using the trapezoidal rule.

### Outcome Assessments

The primary glucoregulatory outcome for this analysis was the Matsuda Index. The trial also included other registered primary outcomes (diet quality assessed by the Healthy Eating Index, microbiome diversity, and serum metabolomics); microbiome and metabolomics findings will be reported in separate manuscripts. While total OGTT AUCs and the Matsuda Index were the registered glucoregulatory measures, segmental OGTT AUCs (0-15, 0-30, 0-60 min) and additional derived indices (HOMA-IR, HOMA-β) are also reported here to capture differential aspects of early-phase and integrated glucose handling. Additional outcomes included anthropometric and body composition measures, blood pressure, EndoPAT measures, fasting lipid profiles, and free-living appetite ratings. Tertiary outcomes included cognitive function measures assessing memory and attention.

### Glucoregulatory Outcomes

#### OGTT Measures

A 2-hour OGTT was conducted at BL and W8. A catheter was inserted and fasting blood was drawn (time 0). Participants then drank 75 g of glucose (Fruit Punch, Fisherbrand™ Glucose Tolerance Test Beverage) over 5 minutes. Additional blood was drawn into SSTs at 15, 30, 60, and 120 minutes after participants finished drinking. Glucose, insulin, and NEFA were measured at each OGTT time point using a YSI 2300D STAT Plus device (Yellow Springs, USA), ELISA (#KAQ1251, Invitrogen, Thermo Fisher Scientific, USA), and a colorimetric assay (NEFA-HR(2) kit, FUJIFILM Wako Diagnostics, USA), respectively. Total and segmental area under the curve (AUC) for glucose, insulin, and NEFA were calculated over the 2-hour period using the trapezoidal rule. Matsuda Index was calculated as 10,000 / √([fasting glucose × fasting insulin] × [mean glucose × mean insulin during OGTT]) (46)

### Fasting Measures

Fasting glucose, insulin, and NEFA values at BL, W4, and W8, measured using the same methods described above for the OGTT. HOMA-IR was calculated as (fasting insulin × fasting glucose) / 405, and HOMA-β as (360 × fasting insulin) / (fasting glucose − 63) (47,48).

### Anthropometric and Body Composition Measures

Height was measured using a wall-mounted stadiometer (SECA 216 Height Measuring Rod, SECA) with participants standing upright, facing forward, with hands on their waist. Weight was measured under light clothing and without bodily attachments using a calibrated electronic scale (Health o meter Professional Scale, Pelstar, model 500KL, USA). Waist and hip circumferences were measured using a standard measuring tape. Waist circumference was recorded at the narrowest point of the torso, whereas hip circumference was recorded at the widest circumference over the hips and gluteal region. All measurements were taken in duplicate and averaged. Waist-hip ratio (WHR) was calculated as average waist divided by average hip circumference.

Body composition was measured with the participant in a reclining position using segmental bioelectrical impedance analysis (BIA; RJL-Systems Quantum-V Segmental, Ref: Q5S, USA). Fat mass, fat-free mass (FFM), Torso fat, and lean dry mass (LDM) were derived from BIA outputs. Fat mass index (FMI) and Fat-free mass index (FFMI) were calculated as fat mass and fat-free mass divided by height squared (kg/m²). All measures were assessed at BL, W4, and W8.

### Cardiovascular Outcomes

Resting systolic (SBP) and diastolic blood pressure (DBP) were measured using an automated sphygmomanometer (OMRON model BP725, Kyoto, Japan) on the non-dominant arm. Reactive hyperemia-peripheral arterial tonometry (RH-PAT) was assessed using an EndoPAT device (EndoPAT 2000, Itamar Medical, Israel), as previously described (49,50). Reactive hyperemia index (RHI) and augmentation index (AI) measures were assessed at BL and W8.

Fasting serum triglycerides (TG; E-BC-K238), total cholesterol (TC; E-BC-K109-M), and HDL-C (E-BC-K221) concentrations were determined using commercially available colorimetric kits (Elabscience, USA). LDL-C was calculated using the Friedewald equation (51): LDL-C = (TC − HDL-C) − (TG/2.2). All measures were assessed at BL, W4, and W8.

### Cognitive Function Outcomes

After the 30-minute OGTT blood draw at BL and W8, memory and attention were assessed. The memory test consisted of a ten-word recall task in which participants were read 10 words and asked to recall as many as possible after two minutes; the outcome was the total number of words correctly recalled (52). Attention was assessed using the standardized d2 Test of Attention (53), which yields three primary scores: Total items processed (TN, a measure of processing quantity), TN minus errors (TNE, a measure of accuracy-adjusted processing), and Concentration Performance (CP, calculated as TN minus errors of commission).

### Statistical Analyses

#### Software

Statistical analyses were conducted in R (version 4.5.2) and JMP Pro (version 19).

### Compliance

Three primary adherence questions were addressed among completers: (1) whether counseling visit attendance differed by intervention group and race, (2) whether PD goal attainment differed by goal type and race across weeks, and (3) whether total study food consumption (days) varied by food type and race. Counseling visit attendance and PD goal attainment were analyzed using binomial generalized linear mixed models with a logit link. For counseling attendance, fixed effects included group (PD, CD), race (Black, White), and the group x race interaction. For PD goal attainment, fixed effects included week (W2, W3, W4, W6, W8), goal type (1, 2, 3), race, and their two– and three-way interactions. Model-estimated probabilities and odds ratios summarized effects. Total days each study food was consumed was analyzed using a linear mixed-effects model with fixed effects for food type, race, and the food x race interaction. Pairwise comparisons were adjusted using the multivariate t (mvt) method (54).

### Food Acceptance and Palatability

Acceptance scores were analyzed as ordinal outcomes using cumulative link mixed models with a logit link; palatability scores were analyzed as continuous outcomes using linear mixed-effects models. Both models included fixed effects for week (W2, W4, W6, W8), food, race, and their two– and three-way interactions, with a random intercept for participant ID to account for repeated measures. Statistically significant fixed effects were further examined using planned contrasts adjusted with the mvt method (54).

### Outcome Analyses

All analyses followed a modified intention-to-treat (mITT) framework including all randomized participants with available outcome data, with missing values addressed via multiple imputation as described below.

Outcomes assessed at baseline and W8 only, including OGTT-derived outcomes (segmental and total glucose, insulin, and NEFA AUC values; Matsuda Index), EndoPAT measures, free-living appetite ratings, and cognitive function, were analyzed using baseline-adjusted linear models (lm function in R (55)). Each model included fixed effects for experimental group (PD, CD), race (Black, White), and the group x race interaction, with the corresponding baseline value as a covariate.

Outcomes assessed at baseline, W4, and W8, including fasting glucose, insulin, NEFA, HOMA-IR, HOMA-β, anthropometric and body composition measures, blood pressure, and fasting lipid profiles, were analyzed using baseline-adjusted linear mixed-effects models (lmer function, lme4 package in R (56)). Fixed effects included group, race, week (W4/W8), and all two– and three-way interactions, with the baseline value as a covariate and a random intercept for participant ID.

Dietary intake outcomes (baseline, W4, W8) were analyzed using the linear mixed-effects approach with additional adjustment for total daily energy intake. Physical activity outcomes (baseline, W4, W8) were analyzed using the linear mixed-effects approach with additional adjustment for total accelerometer wear time. Sleep was analyzed using the linear mixed-effects approach with measures at W1 (baseline), W4/W5 (midpoint), and W8. Sensitivity analyses were also performed for all outcomes additionally adjusting for baseline BMI and food security score.

Estimated marginal means, planned contrasts, and joint tests of model terms were computed using the emmeans package (57). Pairwise comparisons were adjusted for multiple testing using the mvt method (54).

The adequacy of model assumptions was investigated through diagnostic evaluation of model residuals. This included assessment of residual skewness and kurtosis, testing for homogeneity of variance using Levene’s test, and graphical examination of residual patterns and distributions. For variables that failed to meet distributional criteria, Johnson transformation procedures were applied in JMP Pro version 19.0. When analyses based on transformed data led to the same inferential outcomes as analyses based on original-scale data, findings from the original-scale models are reported for interpretability. In cases where transformation affected the statistical interpretation, results from the transformed analyses are presented.

### Missing Value Imputation

Missing observations were handled before statistical analysis using multiple imputation by chained equations, performed with the mice package in R and predictive mean matching as the imputation method (58). The imputation model included sex, age, BMI, food security score, and other timepoint values of each outcome as predictors. Imputations were performed separately for each group x race combination. The required number of imputed datasets was selected according to the von Hippel recommendation, using the largest observed fraction of missingness across variables (59). For each imputed dataset, 50 iterations were run per chain, statistical models were fitted independently, and resulting parameter estimates were combined using Rubin’s rules (60). Pooled model estimates were then used to obtain estimated marginal means, prespecified contrasts, and omnibus tests of model effects with the emmeans package (57). P-values for pairwise comparisons were adjusted for multiple testing using mvt method (54).

## RESULTS

The baseline characteristics of the participants are shown in **Table 1**. Adverse effects were monitored through self-reporting by participants. No adverse effects were reported during the trial.

**Table 1.**
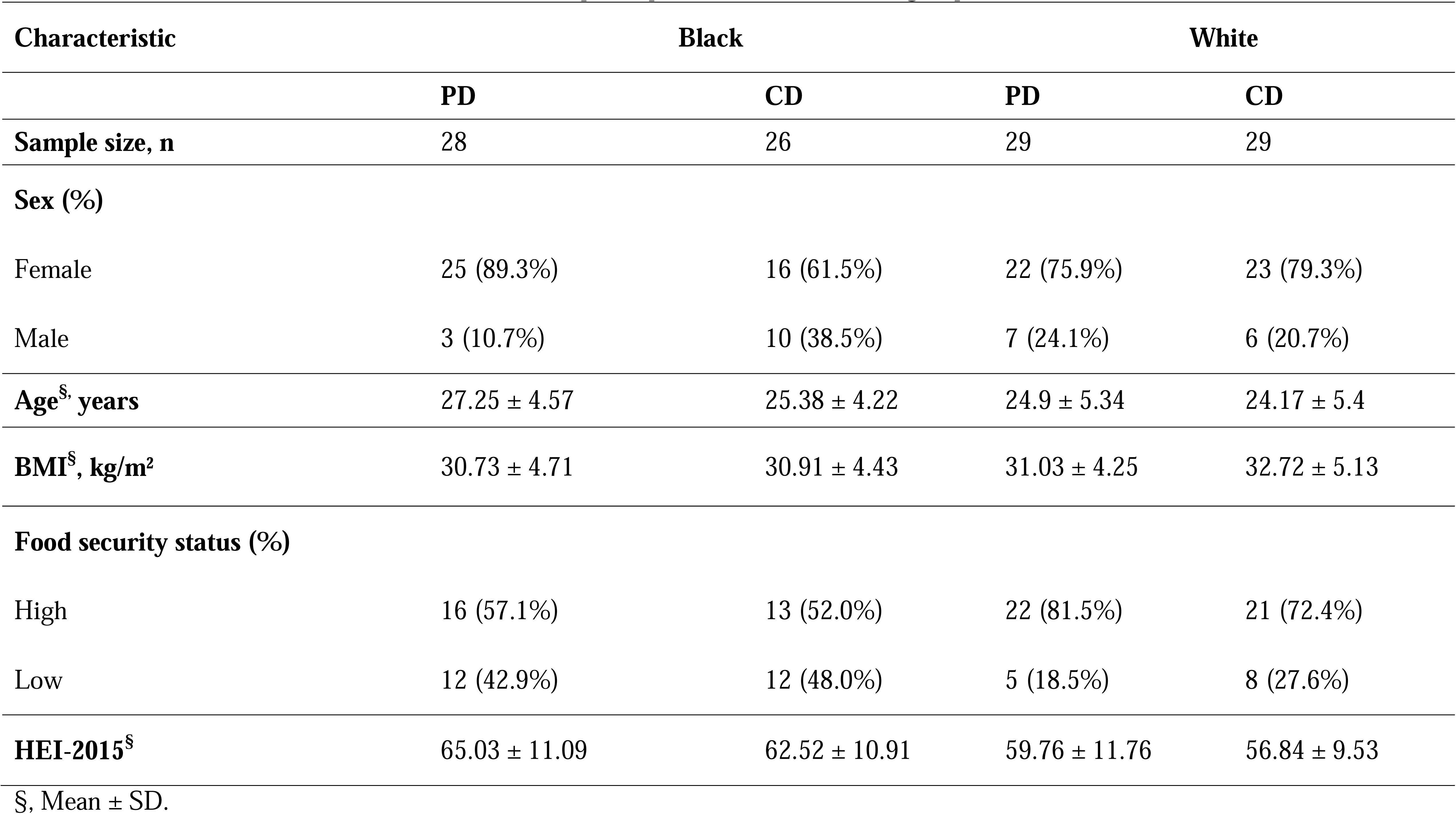
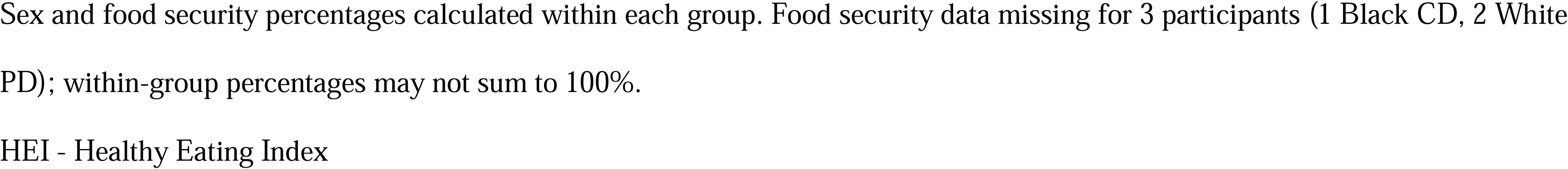
Baseline characteristics of Black and White participants in the PD and CD groups.

### Compliance

#### Counseling visit attendance

Counseling visit attendance differed significantly by group and race (group and race effects, P<0.05, **Supplemental Table 2A**). Attendance was higher in the PD group that received dietary counseling than the CD group that only received counseling at week 1 with food recall visits in the subsequent weeks (83.3% vs. 75.6%, PD vs. CD OR = 1.61, P < 0.05), and lower among Black than White participants (76.3% vs. 82.8%, Black vs. White OR = 0.67, P < 0.05). The group x race interaction was not significant.

### Goal attainment

The overall likelihood of meeting the goals was 74.3% for Goal 1, 77.6% for Goal 2, and 68.1% for Goal 3. In the PD group, there was a statistically significant race x goal effect for goal attainment regardless of week (P<0.05, **Supplemental Table 2B**). Black participants had significantly higher odds of achieving goal 2 (increased unsaturated fat/saturated fat ratio) than White participants (86.7% vs. 64.6%, OR = 3.57, P < 0.05). There were no race effects observed for goal 1 (increased fiber intake) and goal 3 (personalized goal).

### Study food consumption

Mean food consumption for the PD group, expressed as a percentage of the minimum required intake target across the 8-week study (5 days/week; 40 required intake days), was highest for nuts at 91.2%, followed by prunes at 90.5%, carrots at 90.1%, clementines at 89.5%, peppers at 87.3%, and broccoli at 85.1%. Across the overall intervention period, broccoli was consumed less than carrots, clementines, nuts, and prunes (food effect, P<0.05; pairwise food contrasts, all P<0.05, **Supplemental Table 2C**) but not peppers. There were no race or food x race effects observed for total days of consumption.

### Acceptance and Palatability of Foods

Acceptance and palatability scores showed significant food x race interaction effects across all weeks (P<0.05, **Supplemental Table 3**). The acceptance of clementines and prunes compared with walnuts differed by race, with clementines and prunes rated more favorably relative to walnuts among Black participants than among White participants (Clementines-Walnuts: Black vs White, and Prunes-Walnuts: Black vs White, P<0.05). The palatability of clementines compared with walnuts differed by race, with clementines rated more favorably relative to walnuts among Black participants than among White participants (Clementines-Walnuts: Black vs White, P<0.05).

### Dietary Intake

Dietary intake data are shown in **Table 2** and **Supplemental Table 4**. Overall, PD participants had significantly better diet quality than CD participants across all four diet quality indices as reflected in higher HEI-2015, AHEI, DASH, and MED diet scores (group effect, all P<0.05, **Table 2**). PD participants had higher intakes of carbohydrate, total dietary fiber (soluble and insoluble), total monounsaturated and polyunsaturated fats, potassium, magnesium, manganese, dietary folate, vitamin C, vitamin B6, total alpha– and beta-tocopherol, arginine, beta-cryptoxanthin, and total sugars compared to CD (group effect, all P<0.05, **Table 2** and **Supplementary Table 4**). Conversely, PD participants had lower intakes of total dairy, total grains, total protein, added sugars, total saturated fats, conjugated linoleic acid, delta-tocopherol, vitamin B12, vitamin D, sodium, lycopene, and retinol compared to CD (group effect, all P<0.05, **Table 2** and **Supplementary Table 4**). Vitamin D showed a significant week x race interaction, with W4-to-W8 changes varying by race (W4–W8, Black vs. White, P<0.05), increasing among Black participants and decreasing among White participants (**Supplementary Table 4**).

**Table 2.** Self-reported dietary intake from the Diet History Questionnaire III for Black and White participants assigned to the PD and CD groups at Week 4 and Week 8, with adjustment for baseline intake and total energy intake.

| Outcome | Week | PD |  | CD |  | BL-adjusted P-Values |  |  |  |  |  |  |
| --- | --- | --- | --- | --- | --- | --- | --- | --- | --- | --- | --- | --- |
|  |  | Black | White | Black | White | G | R | W | G x R | W x G | W x R | W x G x R |
| Diet quality indices |  |  |  |  |  |  |  |  |  |  |  |  |
| HEI-2015 total score | W4 | 75.91 ± 1.55 | 77.48 ± 1.53 | 62.31 ± 1.84 | 59.84 ± 1.54 | <0.001 | 0.894 | 0.420 | 0.187 | 0.255 | 0.793 | 0.908 |
|  | W8 | 76.06 ± 1.7 | 77.9 ± 1.52 | 60.18 ± 2.52 | 58.39 ± 1.57 |  |  |  |  |  |  |  |
| AHEI total score | W4 | 67.29 ± 1.6 | 63.93 ± 1.6 | 50.41 ± 1.7 | 47.09 ± 1.56 | <0.001 | 0.197 | 0.211 | 0.932 | 0.172 | 0.134 | 0.896 |
|  | W8 | 65.81 ± 1.77 | 65.59 ± 1.57 | 46.5 ± 1.91 | 45.8 ± 1.56 |  |  |  |  |  |  |  |
| Mediterranean (MED) total score | W4 | 5.29 ± 0.25 | 5.43 ± 0.25 | 3.04 ± 0.25 | 2.73 ± 0.26 | <0.001 | 0.838 | 0.737 | 0.633 | 0.473 | 0.384 | 0.429 |
|  | W8 | 5.45 ± 0.28 | 5.61 ± 0.25 | 2.72 ± 0.29 | 2.93 ± 0.25 |  |  |  |  |  |  |  |
| DASH total score | W4 | 27.31 ± 0.77 | 26.91 ± 0.7 | 21.7 ± 0.85 | 20.73 ± 0.71 | <0.001 | 0.884 | 0.754 | 0.589 | 0.319 | 0.212 | 0.925 |
|  | W8 | 27.02 ± 0.84 | 27.86 ± 0.74 | 20.54 ± 0.96 | 20.64 ± 0.75 |  |  |  |  |  |  |  |

| Macronutrients |  |  |  |  |  |  |  |  |  |  |  |  |
| --- | --- | --- | --- | --- | --- | --- | --- | --- | --- | --- | --- | --- |
| **Carbohydrate (g)** | W4 | 175.23 ± 7.51 | 173.62 ± 7.07 | 174.31 ± 10.49 | 148.29 ± 7.56 | **0.015** | 0.335 | 0.394 | 0.272 | 0.272 | 0.165 | 0.450 |
|  | W8 | 173.02 ± 9.25 | 177.53 ± 7.68 | 153.82 ± 9.86 | 148.7 ± 9.83 |  |  |  |  |  |  |  |
| **Protein (g)** | W4 | 50.58 ± 3.53 | 52.44 ± 3.17 | 53.16 ± 3.88 | 66.14 ± 3.6 | **0.024** | 0.211 | 0.955 | 0.279 | 0.635 | 0.149 | 0.241 |
|  | W8 | 52.47 ± 3.79 | 52.75 ± 3.63 | 57.59 ± 4.09 | 59.99 ± 4.29 |  |  |  |  |  |  |  |
| **Total fat (g)** | W4 | 54.11 ± 3 | 59.44 ± 2.9 | 50.23 ± 3.05 | 57.02 ± 3.2 | 0.584 | 0.125 | 0.715 | 0.648 | 0.378 | 0.266 | 0.342 |
|  | W8 | 53.22 ± 3.33 | 58.25 ± 2.97 | 56.86 ± 3.47 | 55.66 ± 3.82 |  |  |  |  |  |  |  |
| **Saturated fat (g)** | W4 | 13.22 ± 1.05 | 14.85 ± 1.02 | 16.45 ± 1.2 | 20.86 ± 1.17 | **<0.001** | **0.037** | 0.776 | 0.560 | 0.437 | 0.208 | 0.208 |
|  | W8 | 12.83 ± 1.14 | 14.44 ± 1.03 | 18.94 ± 1.31 | 19.96 ± 1.3 |  |  |  |  |  |  |  |
| **Monounsaturated fats (g)** | W4 | 23.08 ± 1.47 | 25.34 ± 1.47 | 18.82 ± 1.43 | 19.63 ± 1.43 | **0.001** | 0.436 | 0.856 | 0.284 | 0.540 | 0.527 | 0.552 |
|  | W8 | 22.61 ± 2.09 | 24.89 ± 1.47 | 20.75 ± 1.47 | 19.3 ± 1.52 |  |  |  |  |  |  |  |
| **Polyunsaturated fats (g) †** | W4 | -0.75 ± 0.14 | -0.7 ± 0.13 | -1.23 ± 0.17 | -1.34 ± 0.14 | **0.001** | 0.509 | 0.697 | 0.129 | 0.269 | 0.583 | 0.190 |
|  | W8 | -0.89 ± 0.21 | -0.7 ± 0.14 | -0.91 ± 0.17 | -1.37 ± 0.15 |  |  |  |  |  |  |  |
| **Total dietary fiber (g)** | W4 | 24.14 ± 1.11 | 24.44 ± 1.09 | 13.97 ± 1.17 | 13.32 ± 1.07 | **<0.001** | 0.873 | 0.769 | 0.649 | 0.405 | 0.985 | 0.986 |
|  | W8 | 24.86 ± 1.31 | 25.16 ± 1.14 | 13.62 ± 1.3 | 13.02 ± 1.19 |  |  |  |  |  |  |  |
| **Added sugars (g)** | W4 | 24.62 ± 3.88 | 28.64 ± 3.83 | 42.02 ± 5.03 | 40.58 ± 3.73 | **<0.001** | 0.287 | 0.228 | 0.719 | 0.598 | 0.196 | 0.466 |
|  | W8 | 21.99 ± | 28.38 ± | 34.32 ± | 41.05 ± |  |  |  |  |  |  |  |

|  |  | 3.86 | 3.88 | 4.31 | 3.92 |  |  |  |  |  |  |  |
| --- | --- | --- | --- | --- | --- | --- | --- | --- | --- | --- | --- | --- |
| Micronutrients |  |  |  |  |  |  |  |  |  |  |  |  |
| †Alpha-carotene (µg)<sup>a</sup> | W4 | 0.78 ± 0.1 | 0.58 ± 0.1 | -1.02 ± 0.13 | -0.75 ± 0.1 | <0.001 | 0.283 | 0.464 | **0.024** | 0.690 | 0.348 | 0.716 |
|  | W8 | 0.71 ± 0.1 | 0.69 ± 0.1 | -1 ± 0.15 | -0.63 ± 0.1 |  |  |  |  |  |  |  |
| Beta-carotene (µg)<sup>a</sup> | W4 | 10618.56 ± 564.85 | 8074.47 ± 576.39 | 2084.38 ± 580.22 | 2197.95 ± 528.88 | <0.001 | 0.072 | 0.280 | **0.011** | 0.780 | 0.348 | 0.770 |
|  | W8 | 10674.28 ± 648.16 | 8931.93 ± 602.91 | 2144.17 ± 585.96 | 2674.92 ± 531.91 |  |  |  |  |  |  |  |
| Lutein/zeaxanthin (µg)<sup>a</sup> | W4 | 4232.7 ± 468.38 | 2500.77 ± 412.45 | 1789.2 ± 491.71 | 2061.77 ± 415.54 | <0.001 | 0.053 | 0.255 | **0.002** | 0.422 | 0.831 | 0.482 |
|  | W8 | 4993.68 ± 531.89 | 2777.91 ± 416.86 | 1757.58 ± 454.49 | 2292.84 ± 434.22 |  |  |  |  |  |  |  |
| Sodium (mg) | W4 | 2016.9 ± 123.54 | 1957.74 ± 113.45 | 2165.97 ± 131.45 | 2533.03 ± 130.55 | **0.005** | 0.343 | 1.000 | 0.100 | 0.793 | 0.512 | 0.656 |
|  | W8 | 2053.21 ± 125.75 | 1964.95 ± 121.78 | 2226.71 ± 162.39 | 2428.81 ± 163.93 |  |  |  |  |  |  |  |
| Total alpha-tocopherol (mg) | W4 | 14.4 ± 1.07 | 16.8 ± 1.14 | 8.99 ± 1.12 | 7.46 ± 1.02 | **<0.001** | 0.829 | 0.470 | 0.089 | 0.444 | 0.724 | 0.486 |
|  | W8 | 15.11 ± 1.26 | 16.25 ± 1.1 | 7.88 ± 1.08 | 6.71 ± 1.04 |  |  |  |  |  |  |  |
| Vitamin K (µg)<sup>a</sup> | W4 | 285.74 ± 23.05 | 194.42 ± 21.29 | 105.03 ± 24.29 | 115.9 ± 21.26 | <0.001 | 0.060 | 0.275 | **0.003** | 0.385 | 0.848 | 0.806 |
|  | W8 | 312.61 ± 26.75 | 219.97 ± 21.36 | 102.34 ± 22.31 | 124.86 ± 21.36 |  |  |  |  |  |  |  |

| Food groups |  |  |  |  |  |  |  |  |  |  |  |  |
| --- | --- | --- | --- | --- | --- | --- | --- | --- | --- | --- | --- | --- |
| **Total vegetable cups<sup>a</sup>** | W4 | 2.82 ± 0.14 | 2.42 ± 0.14 | 1.05 ± 0.15 | 1.13 ± 0.13 | <0.001 | 0.335 | 0.198 | **0.047** | 0.465 | 0.669 | 0.841 |
|  | W8 | 2.95 ± 0.16 | 2.58 ± 0.15 | 1.04 ± 0.15 | 1.21 ± 0.14 |  |  |  |  |  |  |  |

BL-adjusted emmeans ± SE are shown for W4 and W8 across Group x Race combinations. †Outcome was Johnson-transformed for analysis; emmeans presented on the original (untransformed) scale where applicable. Repeated measures were analyzed using linear mixed-effects models (fixed effects: G = group, W = week, R = race, GxR = group x race, GxW = group x week, WxR = week x race, WxGxR = week x group x race; random intercept: participant). PD = personalized whole foods group, CD = control group, W4 = week 4, W8 = week 8. Pairwise comparisons used multivariate-t (mvt) adjustment. All analyses followed a modified intention-to-treat (mITT) framework including all randomized participants with valid outcome data, with missing values addressed via multiple imputation.
Significant contrasts: a, PD-CD: Black>White

Although PD improved vegetable-related outcomes relative to CD in both race groups, the PD−CD difference was significantly larger among Black participants than White participants for total vegetable intake, AHEI and DASH vegetable component scores, and vegetable-derived micronutrients including alpha-carotene, beta-carotene, lutein/zeaxanthin, and vitamin K. HEI-2015 total fruits showed the opposite group x race pattern, with the PD−CD difference larger among White participants than Black participants. Cholesterol intake showed a significant week x group x race interaction. Within CD, the W4-to-W8 change varied by race (CD: W4–W8, Black vs. White, P<0.05), increasing among Black participants and decreasing among White participants, **Supplementary Table 4**).

### Physical Activity, Sleep, and Appetite

Physical activity measures such as time spent in sedentary, light, moderate, and vigorous activities, and sleep durations were assessed at BL, W4, and W8, while subjective fullness, hunger, desire to eat, and prospective consumption ratings over the morning, afternoon, evening, and 14-hour periods were assessed at BL and W8. No significant group x race interaction effects were observed for any physical activity, sleep or appetite outcomes (**Supplementary Table 5**). However, PD participants spent significantly less time in light-intensity physical activity than CD participants, and Black participants spent less time in light-intensity activity than White participants (group and race main effects, respectively, P<0.05). Regardless of diet group, Black participants had a lower percentage time in light-intensity activity and higher percentage time in sedentary activity at W4 than White participants. This difference narrowed from W4 to W8, with Black participants showing a greater increase in percentage time in light-intensity activity and reduction in percentage sedentary time relative to White participants over this period (week x race interaction effect P<0.05; W4: Black vs White, P<0.05; W4 to W8 change: Black vs White, P<0.05). Sleep duration demonstrated significant week x race interaction effects (P<0.05) with Black participants having lower sleep duration compared to White participants at W8 (W8: Black vs. White, P<0.05). Morning AUC for fullness and Evening AUC for prospective consumption were higher at W8 for White individuals compared with Black individuals (race effect, P<0.05).

### Glucoregulatory Outcomes

Fasting serum glucose, insulin, and NEFA, along with derived indices (HOMA-IR, HOMA-β), were assessed at BL, W4, and W8 (**Figure 2, Supplemental Table 6**). HOMA-β was significantly lower in PD than CD participants overall (group effect, P<0.05). Moreover, there was a tendency toward a week x group x race interaction for HOMA-IR (P=0.076) and fasting insulin (P=0.064). In a sensitivity analysis additionally adjusting for baseline BMI and food security score, the week x group x race interaction for fasting insulin reached significance (P<0.05) and the corresponding tendency for HOMA-IR remained (P=0.069). For fasting insulin, Black participants in the PD group showed a significant decrease from W4 to W8 (W8 vs. W4 within PD Black, P<0.05), and this W4 to W8 decrease was significantly greater than that in White participants (PD: W4-W8 change: Black vs. White, P<0.05). The PD−CD difference at W8 also differed in direction between races (W8 PD−CD, Black vs. White, P < 0.05), with lower values in PD than CD among Black participants but higher values in PD than CD among White participants. A similar directional pattern was observed for HOMA-IR.

**Figure 2.**
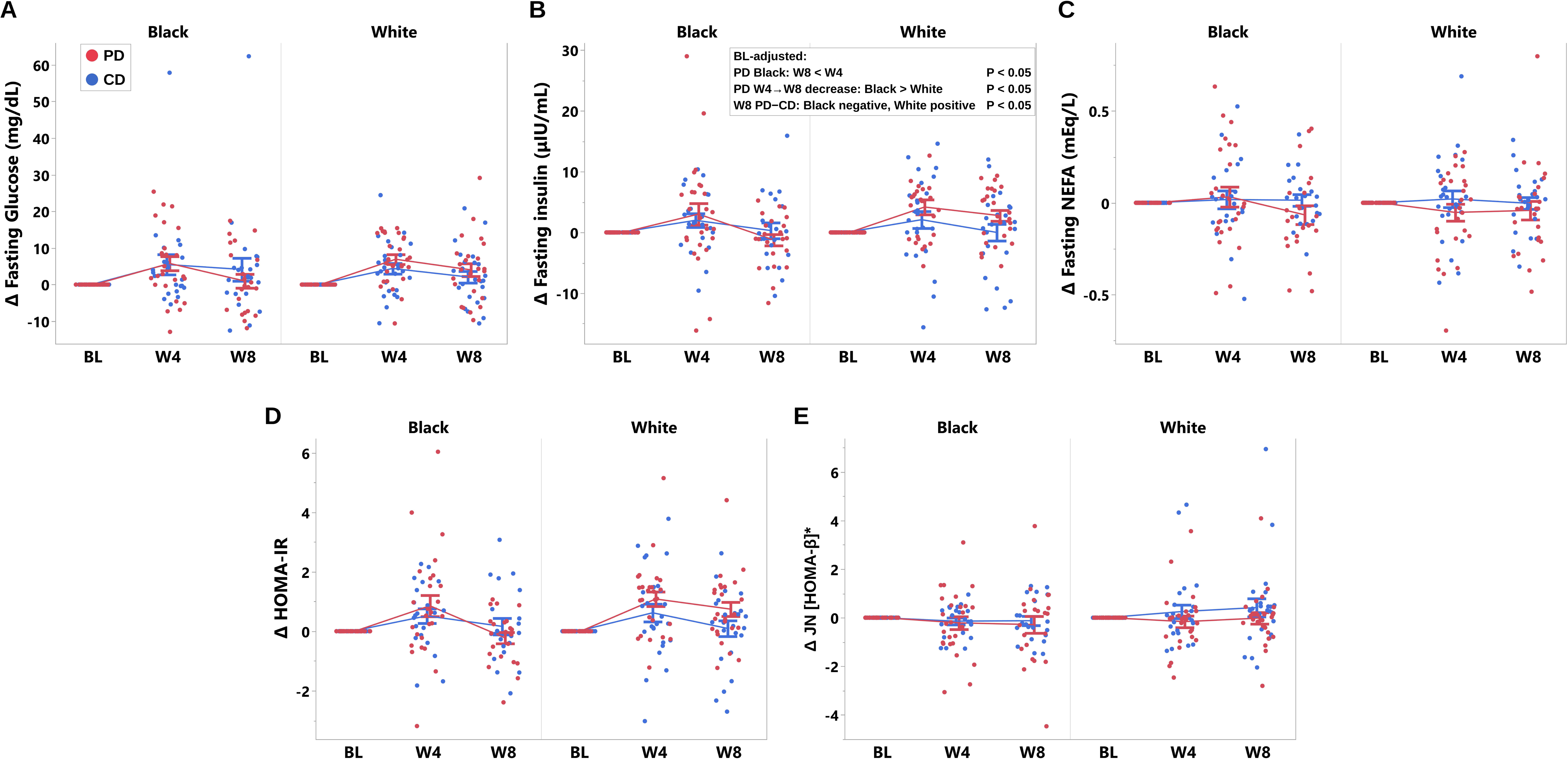
Fasting glucoregulation-associated marker changes from baseline at Week 4 (W4) and Week 8 (W8) for Black and White participants in the PD and CD groups. Change scores (Δ from baseline) are shown for: (A) fasting glucose; (B) fasting insulin; (C) fasting NEFA; (D) HOMA-IR; (E) Δ JN [HOMA-β]*. Baseline is anchored at zero by definition. Individual participant points are jittered horizontally; group means ± SE are connected by lines. Fasting measures were analyzed using baseline-adjusted linear mixed-effects models (fixed effects: G = group, W = week, R = race, G x R = group x race, G x W = group x week, W x R = week x race, W x G x R = week x group x race; random intercept: participant). Pairwise comparisons used multivariate-t (mvt) adjustment. All analyses followed a modified intention-to-treat (mITT) framework including all randomized participants with valid outcome data, with missing values addressed via multiple imputation. In (B), the boxed P-values are post hoc contrasts following a significant Week x Group x Race interaction in the sensitivity analysis additionally adjusted for baseline BMI and food security score. *\**P < 0.05 for the Group main effect (PD < CD). JN denotes Johnson-transformed outcome.

OGTT glucose, insulin, and NEFA time-course curves are presented in **Figure 3**. OGTT-derived outcomes, including segmental and total glucose, insulin, NEFA AUC, and Matsuda Index, were assessed at BL and W8 (**Figure 4, Supplemental Table 6**). Glucose AUC 0-15 min and 0–30 min, reflecting the early glucose response, showed a significant group x race interaction (**Figure 4, Supplemental Table 6**, P<0.05). White participants in the PD group had higher Glucose AUC 0–30 min than Black participants (PD: Black vs. White, P<0.05). The PD−CD difference at W8 also differed in direction between races (W8 PD−CD, Black vs. White, P < 0.05), with lower values in PD than CD among Black participants but higher values in PD than CD among White participants.

**Figure 3.**
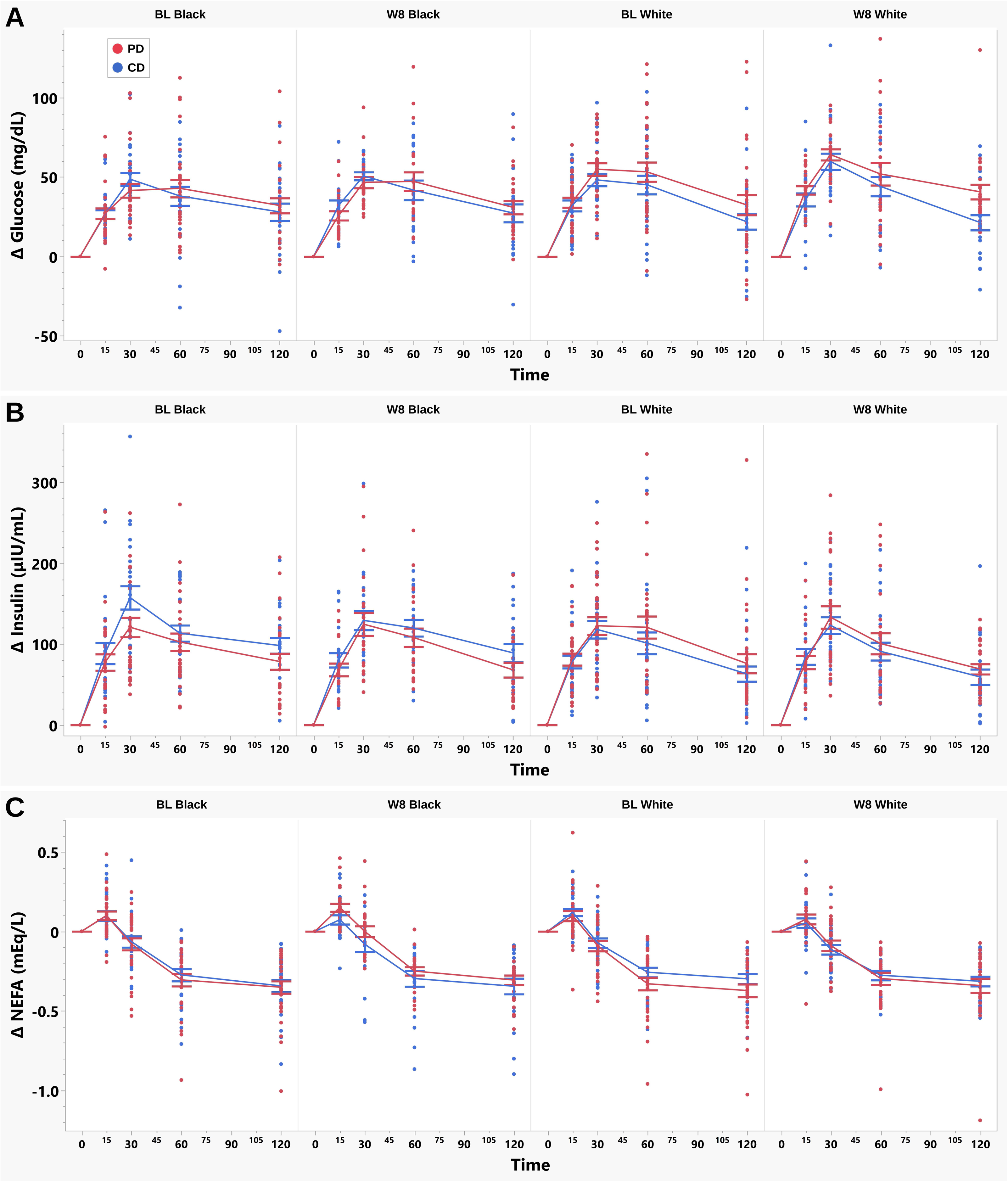
Oral glucose tolerance test (OGTT) time-course responses for glucose, insulin, and NEFA at baseline (BL) and Week 8 (W8) in Black and White participants in the PD and CD groups. (A) Δ Glucose, (B) Δ Insulin, (C) Δ NEFA at 0, 15, 30, 60, and 120 min. Time 0 is anchored at zero by definition. Individual participant points are shown; group means ± SE are connected by lines.

**Figure 4.**
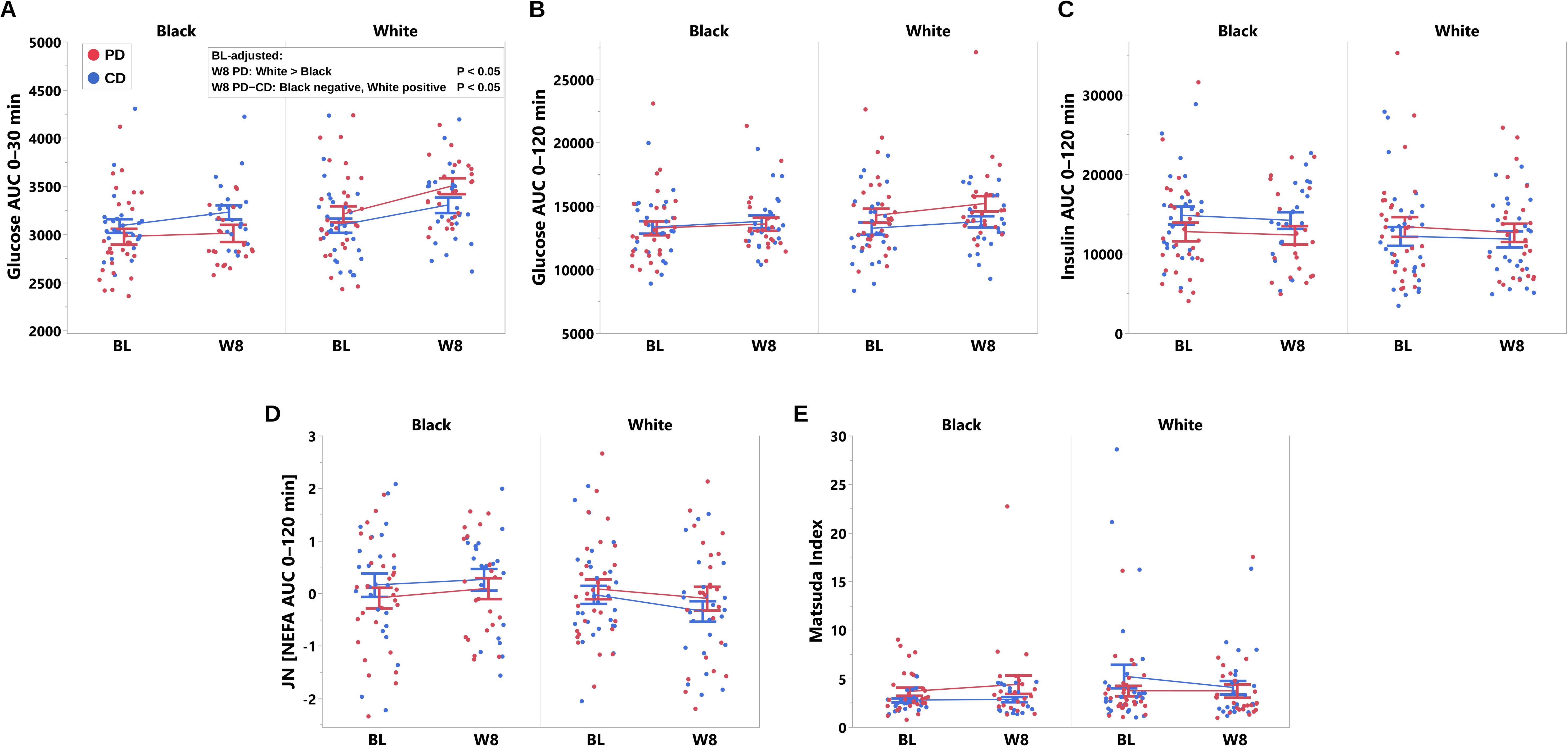
OGTT-derived summary measures at baseline (BL) and Week 8 (W8) in Black and White participants in the PD and CD groups. (A) Glucose AUC 0–30 min; (B) Glucose AUC 0–120 min; (C) Insulin AUC 0–120 min; (D) JN [NEFA AUC 0–120 min]; (E) Matsuda Index. Individual participant points are jittered; group means ± SE are shown. OGTT-derived measures were analyzed using baseline-adjusted linear models (fixed effects: G = group, R = race, G x R = group x race; baseline value as covariate). Pairwise comparisons used multivariate-t (mvt) adjustment. All analyses followed a modified intention-to-treat (mITT) framework including all randomized participants with valid outcome data, with missing values addressed via multiple imputation. In (A), the boxed P-values are post hoc contrasts following a significant Group x Race interaction. JN denotes Johnson-transformed outcome.

After BL adjustment, no statistically significant group or interaction effects were observed for fasting glucose, fasting NEFAs, other segmental or total glucose, insulin, and NEFA AUCs, or Matsuda Index.

### Anthropometric and Body Composition Outcomes

Anthropometric and body composition outcomes were assessed over BL, W4, and W8 (**Table 3**). After BL adjustment, there were no statistically significant main or interaction effects for weight, waist circumference, and waist-hip ratio. Total fat mass, fat mass index, and lean dry mass all showed significant group x race interaction effects (P<0.05); however, the significant interactions for fat mass and fat mass index were attenuated in the sensitivity analysis adjusting for BL BMI and food security score. The interaction for lean dry mass remained significant with White PD participants having lower lean dry mass than White CD participants over the intervention duration (White PD vs. CD, P<0.05), but not Black participants.

**Table 3.** Anthropometric, body composition, and cardiovascular outcomes for Black and White participants assigned to the PD and CD groups at Week 4 and Week 8, with adjustment for baseline values.

| Outcome | Week | PD |  | CD |  | BL-adjusted P-Values |  |  |  |  |  |  |
| --- | --- | --- | --- | --- | --- | --- | --- | --- | --- | --- | --- | --- |
|  |  | Black | White | Black | White | G | R | W | G x R | G x W | W x R | W x G x R |
| Anthropometric & body composition outcomes |  |  |  |  |  |  |  |  |  |  |  |  |
| Weight (kg) | W4 | 87.73 ± | 87.81 ± | 88.68 ± | 88.05 ± | 0.155 | 0.524 | 0.827 | 0.142 | 0.302 | 0.232 | 0.274 |
|  |  | 0.79 | 0.85 | 0.88 | 0.94 |  |  |  |  |  |  |  |
|  | W8 | 85.6 ±<br>1.02 | 88.47 ±<br>0.88 | 89.12 ±<br>0.92 | 88.59 ±<br>1.25 |  |  |  |  |  |  |  |
| **Waist**<br>**circumference**<br>**(cm)** | W4 | 89.72 ±<br>0.63 | 90.4 ±<br>0.63 | 90.07 ±<br>0.69 | 89.66 ±<br>0.65 | 0.377 | 0.275 | 0.603 | 0.155 | 0.088 | 0.239 | 0.58 |
|  | W8 | 88.59 ±<br>0.76 | 90.63 ±<br>0.65 | 90.73 ±<br>0.68 | 90.79 ±<br>0.68 |  |  |  |  |  |  |  |
| **Waist-Hip**<br>**Ratio (WHR)** | W4 | 0.79 ±<br>0.01 | 0.79 ±<br>0.01 | 0.79 ±<br>0.01 | 0.79 ±<br>0.01 | 0.808 | 0.400 | 0.569 | 0.683 | 0.623 | 0.347 | 0.45 |
|  | W8 | 0.79 ±<br>0.01 | 0.79 ±<br>0.01 | 0.78 ±<br>0.01 | 0.79 ±<br>0.01 |  |  |  |  |  |  |  |
| **‡ Fat mass (kg)** | W4 | 33.45 ±<br>0.63 | 33.23 ±<br>0.62 | 33.72 ±<br>0.64 | 32.67 ±<br>0.65 | 0.650 | 0.845 | 0.278 | 0.140 | 0.237 | 0.150 | 0.188 |
|  | W8 | 31.68 ±<br>0.93 | 33.38 ±<br>0.62 | 32.75 ±<br>0.69 | 32.75 ±<br>0.69 |  |  |  |  |  |  |  |
| **Fat mass % (%**<br>**body weight)** | W4 | 37.96 ±<br>0.69 | 38.07 ±<br>0.68 | 37.83 ±<br>0.72 | 37.74 ±<br>0.66 | 0.787 | 0.507 | 0.094 | 0.326 | 0.199 | 0.192 | 0.076 |
|  | W8 | 36.04 ±<br>0.86 | 38.07 ±<br>0.68 | 37.82 ±<br>0.71 | 37.45 ±<br>0.68 |  |  |  |  |  |  |  |
| **Fat-free mass**<br>**(FFM, kg)** | W4 | 53.93 ±<br>0.61 | 53.92 ±<br>0.6 | 54.46 ±<br>0.67 | 54.22 ±<br>0.62 | 0.393 | 0.834 | 0.174 | 0.804 | 0.792 | 0.989 | 0.925 |
|  | W8 | 54.22 ±<br>0.62 | 54.28 ±<br>0.6 | 54.95 ±<br>0.67 | 54.67 ±<br>0.69 |  |  |  |  |  |  |  |
| **Fat-free mass**<br>**% (% body**<br>**weight)** | W4 | 62.04 ±<br>0.69 | 61.96 ±<br>0.67 | 62.19 ±<br>0.72 | 62.23 ±<br>0.65 | 0.758 | 0.494 | 0.078 | 0.325 | 0.167 | 0.178 | 0.065 |
|  | W8 | 63.95 ±<br>0.76 | 61.94 ±<br>0.67 | 62.18 ±<br>0.7 | 62.51 ±<br>0.68 |  |  |  |  |  |  |  |
| **‡ Fat mass**<br>**index (FMI,** | W4 | 12.11 ±<br>0.24 | 12.11 ±<br>0.25 | 12.32 ±<br>0.26 | 11.72 ±<br>0.25 | 0.990 | 0.639 | 0.413 | 0.051 | 0.472 | 0.175 | 0.286 |
| **kg/m<sup>2</sup>)** |  |  |  |  |  |  |  |  |  |  |  |  |
|  | W8 | 11.56 ±<br>0.33 | 12.12.24<br>± 0.25 | 12.26 ±<br>0.28 | 11.73 ±<br>0.26 |  |  |  |  |  |  |  |
| **Lean dry mass**<br>**(LDM, kg) <sup>a</sup>** | W4 | 15.2 ±<br>0.24 | 14.97 ±<br>0.24 | 15.2 ±<br>0.26 | 15.84 ±<br>0.26 | 0.092 | 0.737 | 0.739 | **0.036** | 0.642 | 0.339 | 0.721 |
|  | W8 | 15.47 ±<br>0.27 | 14.89 ±<br>0.24 | 15.27 ±<br>0.27 | 15.74 ±<br>0.29 |  |  |  |  |  |  |  |
| **Torso fat %** | W4 | 21.57 ±<br>0.48 | 21.24 ±<br>0.46 | 21.4 ±<br>0.49 | 21.35 ±<br>0.48 | 0.49 | 0.792 | **0.011** | 0.53 | 0.144 | 0.174 | 0.069 |
|  | W8 | 19.94 ±<br>0.57 | 21.08 ±<br>0.49 | 21.28 ±<br>0.48 | 21.01 ±<br>0.52 |  |  |  |  |  |  |  |
| **Cardiovascular outcomes** |  |  |  |  |  |  |  |  |  |  |  |  |
| **Diastolic blood**<br>**pressure (DBP)** | W4 | 78.71 ±<br>1.05 | 76.24 ±<br>1.07 | 77.3 ±<br>1.11 | 76.85 ±<br>1.09 | 0.708 | 0.179 | 0.587 | 0.459 | 0.119 | 0.741 | 0.564 |
|  | W8 | 77.23 ±<br>1.1 | 75.65 ±<br>1.06 | 77.92 ±<br>1.13 | 77.24 ±<br>1.14 |  |  |  |  |  |  |  |
| **Systolic blood**<br>**pressure (SBP)** | W4 | 118.73<br>± 1.42 | 116.37 ±<br>1.39 | 119 ±<br>1.55 | 115.32 ±<br>1.43 | 0.407 | 0.257 | 0.055 | 0.876 | 0.15 | 0.072 | 0.604 |
|  | W8 | 117.95<br>± 1.53 | 118.02 ±<br>1.42 | 120.05 ±<br>1.69 | 120.69 ±<br>1.6 |  |  |  |  |  |  |  |
| **Total**<br>**cholesterol (TC)**<br>**(mg/dL)** | W4 | 202.27<br>± 9.18 | 200.3 ±<br>8.01 | 196.29 ±<br>8.58 | 190.65 ±<br>8.18 | 0.392 | 0.690 | 0.846 | 0.942 | 0.711 | 0.864 | 0.788 |
|  | W8 | 199.92<br>± 8.8 | 196.97 ±<br>8.05 | 195.03 ±<br>8.57 | 193.75 ±<br>8.79 |  |  |  |  |  |  |  |
| **LDL-C (mg/dL)** | W4 | 136.5 ±<br>7.37 | 130.89 ±<br>6.91 | 130.89 ±<br>7.45 | 123.76 ±<br>6.78 | 0.543 | 0.454 | 0.867 | 0.833 | 0.487 | 0.700 | 0.647 |
|  | W8 | 133.23<br>± 7.55 | 126.89 ±<br>6.97 | 129.49 ±<br>7.47 | 129.54 ±<br>7.33 |  |  |  |  |  |  |  |
| **HDL-C (mg/dL)** | W4 | 52.53 ±<br>1.47 | 52.27 ±<br>1.34 | 52.3 ±<br>1.43 | 51.23 ±<br>1.33 | 0.369 | 0.230 | 0.460 | 0.621 | 0.534 | 0.200 | 0.801 |
|  | W8 | 53.21 ±<br>1.62 | 51.47 ±<br>1.34 | 52.41 ±<br>1.45 | 49.12 ±<br>1.37 |  |  |  |  |  |  |  |
| **Triglycerides**<br>**(mg/dL) †** | W4 | -0.21 ±<br>0.16 | 0.32 ±<br>0.15 | -0.14 ±<br>0.16 | 0.23 ±<br>0.15 | 0.459 | **0.007** | 0.923 | 0.623 | 0.477 | 0.350 | 0.825 |
|  | W8 | -0.03 ±<br>0.16 | 0.27 ±<br>0.15 | -0.15 ±<br>0.16 | 0.08 ±<br>0.16 |  |  |  |  |  |  |  |
| **Augmentation**<br>**index (AI) ^** | W8 | -3.61 ±<br>2.05 | -2.86 ±<br>1.87 | -4.03 ±<br>2.37 | -5.87 ±<br>1.89 | 0.399 | 0.798 | — | 0.523 | — | — | — |
| **Reactive**<br>**hyperemia**<br>**index (RHI) ^** | W8 | 2.09 ±<br>0.12 | 2.24 ±<br>0.13 | 2.07 ±<br>0.13 | 2.32 ±<br>0.13 | 0.816 | 0.124 | — | 0.696 | — | — | — |

BL-adjusted emmeans ± SE are shown for W4 and W8 across Group x Race combinations. †Outcome was Johnson-transformed;
transformed-scale emmeans shown. Repeated measures were analyzed using linear mixed-effects models (fixed effects: G = group, W = week, R = race, G x R = group x race, G x W = group x week, W x R = week x race, W x G x R = week x group x race; random intercept: participant). PD = personalized whole foods group, CD = control group, W4 = week 4, W8 = week 8. ^ Outcome was measured at baseline and W8 only. Pairwise comparisons used multivariate-t (mvt) adjustment. All analyses followed a modified intention-to-treat (mITT) framework including all randomized participants with valid outcome data, with missing values addressed via multiple imputation.
‡ P-values and emmeans shown are from sensitivity analysis additionally adjusting for baseline BMI and food security score; the Group x Race interaction was significant ( $P < 0.05$ ) in the main analysis but attenuated ( $P \geq 0.05$ ) in the sensitivity analysis.
Significant contrasts: a, White: CD > PD

### Cardiovascular Outcomes

Systolic and diastolic blood pressures, and serum markers of LDL-C, HDL-C, total cholesterol, and triglycerides were assessed at BL, W4, and W8 (**Table 3**). EndoPAT measures including RHI and AI, were assessed at BL and W8. After BL adjustment, there were no statistically significant main or interaction effects for any cardiovascular outcomes except for triglycerides that were significantly lower in Black participants than White participants overall (race effect, P<0.05).

### Cognitive Function Outcomes

Immediate memory and attention outcomes were assessed at BL and W8 (**Figure 5, Supplemental Table 7**). After BL adjustment, concentration performance (CP) scores were significantly higher in PD participants than CD participants (group effect, P<0.05). Measures of processing quantity (TN) and accuracy-adjusting processing (TNE) scores were significantly higher in PD participants than CD participants overall (group effect, P<0.05) with the PD vs. CD difference being larger in Black participants than White participants (group x race effect, P<0.05). No significant main or interaction effects were found for memory outcomes.

**Figure 5.**
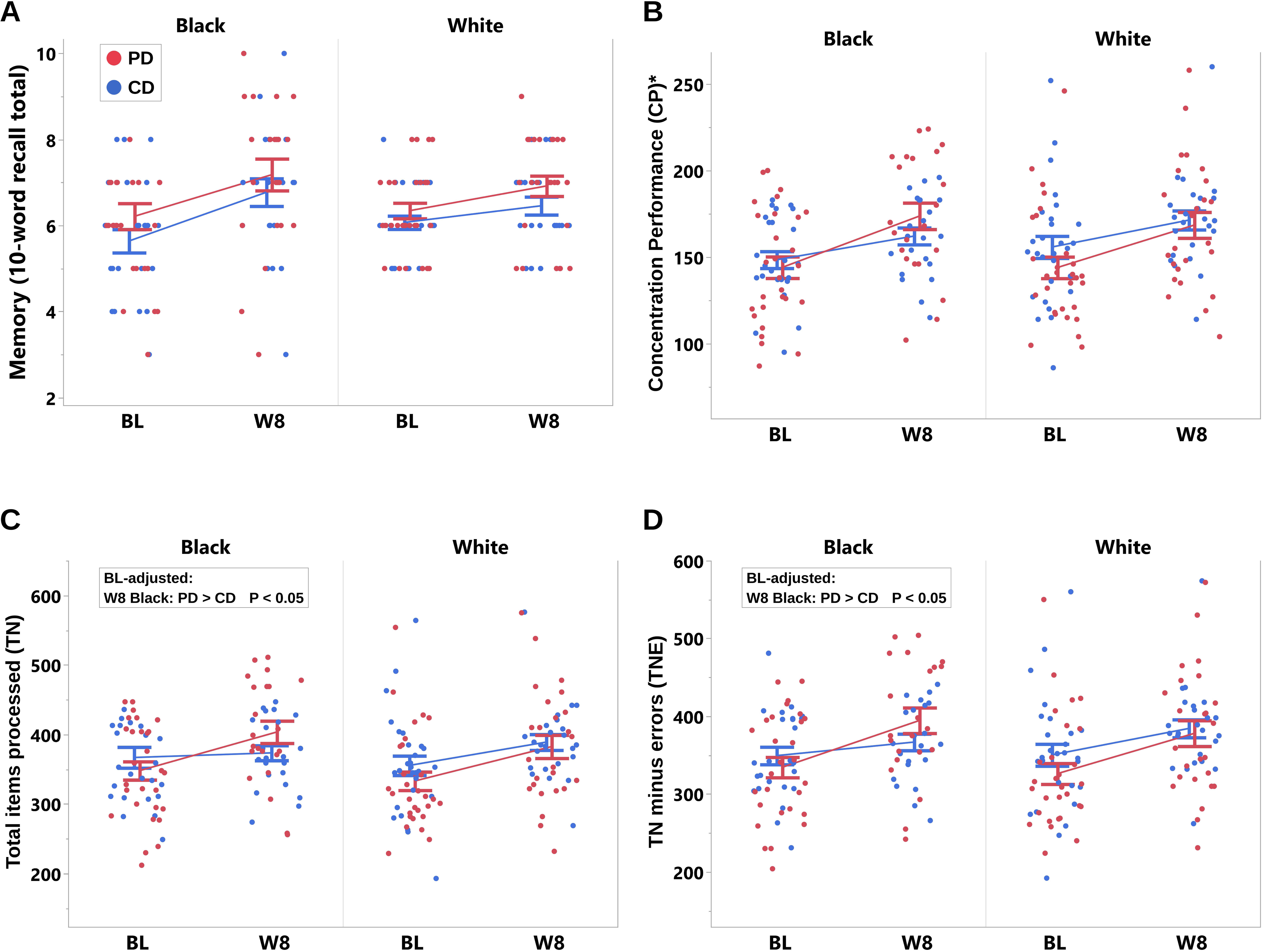
Cognitive function outcomes at baseline (BL) and Week 8 (W8) for Black and White participants in the PD and CD groups. (A) Memory, scored as the total number of words correctly recalled on the 10-word recall task. (B) Concentration Performance (CP)* from the d2 Test of Attention. (C) Total items processed (TN) from the d2 Test of Attention. (D) TN minus errors (TNE) from the d2 Test of Attention. Non-transformed values are presented. Individual participant points are jittered; group means ± SE are connected by lines. Outcomes were analyzed using baseline-adjusted linear models (fixed effects: G = group, R = race, G x R = group x race; baseline value as covariate). Pairwise comparisons used multivariate-t (mvt) adjustment. All analyses followed a modified intention-to-treat (mITT) framework including all randomized participants with valid outcome data, with missing values addressed via multiple imputation. In (C) and (D), the boxed P-values are post hoc contrasts following a significant Group x Race interaction. *P < 0.05 for the Group main effect (PD > CD).

## DISCUSSION

In this 8-week trial of a personalized whole-food intervention versus conventional dietary advice in young Black and White adults with overweight or obesity, the intervention did not improve the primary outcome of whole-body insulin sensitivity as measured by Matsuda Index. However, it reduced beta-cell function index, HOMA-β, and improved concentration performance compared with conventional dietary advice. Intervention-induced improvements were larger in Black participants for intakes of vegetables and associated micronutrients, and attention measures reflecting processing speed and accuracy. Race-differential intervention effects were also observed for fasting insulin and initial glucose responses to OGTT, and body composition measures, although fat mass effects were attenuated after adjusting for BMI and food security status. The remaining glucoregulatory, cardiovascular and appetite outcomes did not differ between groups.

The intervention’s effectiveness was reflected in alignment with healthy dietary patterns such as DASH and MED and achieved through strong adherence with food consumption (85% to 91%) and moderate to strong dietary goal attainment (68% to 78%). Although overall diet quality indices improved comparably between Black and White participants, Black participants were more likely to achieve the unsaturated fat goal and had greater intakes of total vegetables, and micronutrients including alpha– and beta-carotene, lutein and zeaxanthin, and vitamin K. In contrast, prior behavioral weight loss and dietary counseling trials that did not directly provide food have reported modest decreases in fat intake and less pronounced increases in fruit and vegetable intake among African American than non-African American participants (61). In the present study, the greater improvements in specific dietary components observed among Black participants suggest that direct food provision and counseling may help overcome the adherence gap typically observed when dietary interventions rely on education alone.

The intervention modestly reduced HOMA-β, an index of basal β-cell secretory output relative to fasting glucose (62). In adults with overweight or obesity, fasting hyperinsulinemia is a well-established feature of compensatory β-cell secretion in the setting of insulin resistance (63). In this context, a modest reduction in HOMA-β with stable fasting glucose may reflect reduced compensatory insulin secretion rather than declining β-cell secretory capacity, particularly in young adults without overt β-cell dysfunction (48). In the present study, the intervention-induced reduction in fasting insulin, especially during the latter half of the 8-week intervention, along with a tendency toward lower HOMA-IR among Black relative to White participants, may also have clinical relevance because Black adults in the United States have higher fasting insulin concentrations (64) and lower insulin sensitivity than their White counterparts (4). Similar directional findings have been reported broadly for prior whole-food interventions, including a 12-week avocado study that noted a tendency for lower fasting insulin relative to control without changes in Matsuda Index among adults with overweight or obesity and insulin resistance (65), and a mixed-nut intervention that reduced fasting insulin and HOMA-IR among adults with metabolic syndrome (66), although neither trial examined or was designed to test differences by race.

In contrast to the effects observed on fasting-state measures, the intervention did not change the Matsuda Index, a measure of whole-body insulin sensitivity (46). While fasting indices primarily reflect hepatic insulin sensitivity, the Matsuda Index integrates fasting and dynamic OGTT glucose and insulin responses to reflect both hepatic and peripheral contributions (67,68). Reported test-retest coefficients of variation for this index are 18 to 26 percent in adults with normal or impaired fasting glucose (69), indicating substantial inter-individual variability that may reflect factors beyond the dietary intervention, including baseline glucoregulatory phenotype. Mixed Matsuda Index responses have been reported across dietary and lifestyle trials in adults with overweight or obesity, including null findings in a 6-week eucaloric controlled-feeding trial comparing diets with 81 percent versus 0 percent energy from ultraprocessed food in midlife adults (70), a 6-month beverage substitution trial in overweight and obese adults (71), and a 7.5-week sleep-aligned overnight fasting trial in middle-aged and older adults with overweight or obesity that nonetheless improved 30-minute insulinogenic index and OGTT glucose (72). This pattern reflects how intervention effects on the early phases of the OGTT can occur without changing the integrated Matsuda Index.

The present trial supports this by showing race-differential changes in early-phase glucose handling, with the intervention maintaining stability in 0 to 15 and 0 to 30 minute glucose AUC in Black participants compared with the conventional dietary counseling group and with White participants in either arm. During an oral glucose tolerance test, the early glucose rise reflects two glucose sources entering the circulation, glucose absorbed from the gut, and glucose released by the liver through endogenous glucose production. First-phase insulin secretion acts at the liver to suppress endogenous glucose production (73), limiting the hepatic glucose added to the gut-derived load. The degree of suppression depends on two separate factors, the amount of first-phase insulin secreted, and the responsiveness of the liver to that insulin. Because first-phase insulin secretion (insulin AUC 0-30 min) was similar between Black and White participants in the PD group, the race-differential pattern in early-phase glucose AUC cannot be attributed to differential insulin output. The difference must therefore arise at the level of hepatic responsiveness to first-phase insulin, a site where greater hepatic insulin resistance in Black compared with White adults has been hypothesized (67).

However, this interpretation is inferential because the present trial used OGTT-derived indices rather than hyperinsulinemic-euglycemic clamp with isotopic tracer methods, which would be required to directly partition hepatic and peripheral insulin action. Altogether, these findings suggest that fasting measures and early-phase glucose handling can shift independently of the integrated OGTT response (74) and may be differentially influenced by diet-population interactions.

Beyond effects on glucoregulatory outcomes, the intervention improved concentration performance across both race groups, with larger gains in processing speed and accuracy among Black than White participants. As indicated by a recent systematic review of RCTs, healthy dietary patterns, foods, and dietary supplements rich in unsaturated fats, carotenoids, polyphenols, and B vitamins have the potential to improve cognitive domains including attention, concentration, and psychomotor speed (75). Consistent with the present study’s findings, memory (76–78), attention (76,78), and processing speed (77,79) improvements have been reported across studies of high-oleic peanuts, almonds, MIND diet, and lutein and zeaxanthin interventions in adults with overweight or obesity, prediabetes, or healthy adults.

However, direct evidence on dietary intervention effects on cognitive function in Black adults is limited, although in a population analysis of US adults, dietary lutein and zeaxanthin intake was positively associated with cognitive function (digital substitution score test) across racial groups, with the association appearing more pronounced among Black participants (80). Hence, the race-differential improvement in processing speed and accuracy observed in the present trial parallels the race-differential increase in vegetable-derived carotenoids, lutein, and zeaxanthin in the dietary data, providing a coherent dietary-cognitive link. However, the tertiary nature of these outcomes precludes strong mechanistic claims and warrants confirmation in trials specifically designed for cognitive endpoints.

The strengths of this trial include *a priori* race-stratified randomization, a personalized whole-food intervention with tracked goal attainment, concurrent multi-domain outcome assessment, and adjustment for food security score in sensitivity analyses, a structural determinant of dietary behavior with limited incorporation in clinical dietary intervention studies. Some limitations warrant consideration. The 8-week duration may have been insufficient to detect changes in cardiovascular measures. We also did not use the gold-standard direct measurement of hepatic and peripheral insulin sensitivity using the hyperinsulinemic-euglycemic clamp and tracer methodology in this design. However, surrogate insulin indices derived by OGTT have demonstrated strong association with clamp measurements (46). Additionally, body composition was assessed by bioelectrical impedance analysis, a method with lower precision relative to dual-energy X-ray absorptiometry or MRI and uses equations that may perform differently across racial groups, constraining accurate compartmental interpretation of the race-differential lean dry mass effect that remained significant in sensitivity analyses.

In conclusion, this study provides new evidence that this personalized whole-food intervention produced differential effects in fasting insulin and early-phase glucose handling, and greater benefits in attention, in Black compared with White young adults with overweight or obesity during weight maintenance. Several analytic directions follow from these findings. Complier-stratified analyses would refine effect estimates across groups and races. Untargeted metabolomic profiling of banked serum and stool samples could identify objective dietary biomarkers of the dietary intervention as well as early molecular signatures underlying the OGTT and fasting race-differential responses that precede detectable changes in standard clinical indicators. Moreover, to determine which dietary components, individually or in clusters, drive the greatest changes in glucoregulatory and cognitive outcomes, cluster-based or univariate analyses can be conducted in parallel. In future studies, postprandial assessment using mixed-meal challenges, which more closely approximate habitual food intake than the glucose-only OGTT, could test whether the glucoregulatory effects observed here translate to or are strengthened under mixed-macronutrient meal conditions, complementing the OGTT, which remains the standard for clinical interpretation.

## Acknowledgements

JD designed research; JD, OA conducted research; JD, OA analyzed data; JD led the writing of the first draft, with OA contributing to the introduction, methods, and initial drafts, and ER contributing to results presentation and interpretation. All authors revised the paper. JD had primary responsibility for final content. All authors read and approved of the final manuscript. Computational analyses for this work were supported by the high-performance computing infrastructure provided by Research Support Solutions in the Division of Information Technology at the University of Missouri-Columbia (https://doi.org/10.32469/10355/97710).

## Data Availability

The dataset supporting this manuscript will be available from the authors upon reasonable request after completion of all planned study analyses and associated publications.

## Declaration of Generative AI and AI-assisted technologies in the writing process

During the preparation of this work the authors used ChatGPT (5.5) and Claude Opus 4.7 for editing and formatting purposes only. After using this tool/service, the authors reviewed and edited the content as needed and take full responsibility for the content of the publication.

## Abbreviations

AHEI: Alternative Healthy Eating Index
AUC: Area under the curve
BL: Baseline
BMI: Body mass index
CD: Conventional dietary advice
CP: Concentration performance
DASH: Dietary Approaches to Stop Hypertension
HEI: Healthy Eating Index
HOMA-β: Homeostatic Model Assessment of β-cell function
HOMA-IR: Homeostatic Model Assessment of Insulin Resistance
MED: Mediterranean diet
NEFA: Non-esterified fatty acids
OGTT: Oral glucose tolerance test
PD: Personalized whole-food diet
TN: Total items processed
TNE: Total items processed minus errors
W4: Week 4
W8: Week 8

