## Supplemental Tables for "A Personalized Whole-Food Diet Differentially Modulates Glucoregulatory and Cognitive Responses Compared With Conventional Dietary Counseling in Young Black and White Adults With Overweight or Obesity: An 8-Week Randomized Controlled Trial"

**Supplemental Table 1:** Daily quantities and macronutrient composition of study foods provided to participants in PD group

| **Food item** | **Serving size per participant per day** | **Energy (kcal)** | **Protein (g)** | **Total fat (g)** | **Carbohydrate, by difference (g)** | **Fiber, total dietary (g)** | **Saturated fats (g)** | **Monounsaturated fats (g)** | **Polyunsaturated fats (g)** |
| --- | --- | --- | --- | --- | --- | --- | --- | --- | --- |
| **Nuts (total 1.5 oz)** | | | | | | | | | |
| Walnuts,shelled, raw (Sam’s Club Member's Mark) | 0.75 oz | 144.36 | 3.04 | 13.67 | 3.04 | 1.51 | 1.14 | 1.9 | 9.87 |
| Almonds, whole, raw (Sam’s Club Member's Mark) | 0.75 oz | 127.49 | 4.5 | 10.5 | 4.5 | 3 | 0.75 | 6.75 | 2.63 |
| **Fruits (total 2 servings)** | | | | | | | | | |
| Dried Prunes, pitted, no added sugar (Sam’s Club Member's Mark) | 3.00 oz | 207.4 | 1.89 | 0 | 54.74 | 5.7 | 0 | 0 | 0 |
| Clementines (raw, without peel) | 180.00 g | 95.4 | 1.46 | 0.56 | 24.01 | 3.24 | 0.07 | 0.11 | 0.12 |
| **Vegetables (total 3 servings)** | | | | | | | | | |
| Baby carrots | 120.00 g | 42 | 0.77 | 0.16 | 9.89 | 3.48 | 0.03 | 0.01 | 0.08 |
| Broccoli florets | 100.00 g | 35 | 3.53 | 0 | 4.71 | 2.4 | 0 | 0 | 0 |
| Mixed sweet peppers | 110.00 g | 24 | 1.18 | 0 | 5.88 | 1.2 | 0 | 0 | 0 |

Nutrient values were calculated using a combination of the USDA FoodData Central nutrient database and branded product Nutrition Facts labels, as applicable. Energy values may not exactly match energy calculated from protein, carbohydrate, and fat using standard 4/4/9 kcal/g factors because of rounding, fiber content, and database- or label-specific energy calculations.

**Supplemental Table 2.** Compliance outcomes by intervention group and race: A) counseling visit attendance, B) dietary goal attainment and C) study food consumption.

**A. Counseling visit attendance percentage by intervention group and race**

|  | **Group** | | **Race** | | **P-values** |
| --- | --- | --- | --- | --- | --- |
|  | **PD** | **CD** | **Black** | **White** |  |
| Attendance rate, % (mean ± SE) | 83.32 ± 2.04 | 75.61 ± 2.40 | 76.26 ± 2.43 | 82.83 ± 2.03 | **Group: <0.05**  **Race: <0.05**  Group x Race: 0.230 |
| Odds ratio (95% CI) | PD vs. CD:  1.61 (1.10–2.37) | | Black vs. White  0.67 (0.45–0.98) | |  |

**B. Dietary goal attainment percentage in the PD group by race**

| **Goal** | **Black**  **Mean ± SE (%)** | **White**  **Mean ± SE (%)** | **Black vs. White**  **OR (95% CI)** | **P-values** |
| --- | --- | --- | --- | --- |
| Goal 1: Increased dietary fiber | 71.10 ± 5.44 | 77.18 ± 4.42 | 0.73 (0.26–2.06) | Week: 0.122  Goal: 0.125  Race: 0.098  Week x Goal: 0.365  Week x Race: 0.118  **Race x Goal: <0.05**  Week x Race x Goal:0.585 |
| Goal 2: Increased unsaturated/saturated fat ratio* | 86.73 ± 3.87 | 64.64 ± 5.49 | 3.57 (1.10–11.65) |  |
| Goal 3: Personalized goal | 72.33 ± 5.70 | 63.53 ± 5.41 | 1.50 (0.52–4.30) |  |

**C. Study food consumption percentage in the PD group**

| **Study food** | **Mean ± SD (%)** | **P-values** |
| --- | --- | --- |
| Broccoli^b^ | 85.11 ± 15.64 | **Food: <0.05**  Race: 0.875  Food x Race: 0.872 |
| Carrots^a^ | 90.05 ± 12.22 |  |
| Clementines^a^ | 89.52 ± 14.07 |  |
| Nuts^a^ | 91.22 ± 12.01 |  |
| Peppers^ab^ | 87.34 ± 16.60 |  |
| Prunes^a^ | 90.48 ± 12.57 |  |

Panel A counseling visit attendance was analyzed using a binomial generalized linear mixed model with a logit link, using all scheduled counseling visit records during the intervention period as attended/not attended. The model included intervention group, race, and their interaction.

Panel B PD goal attainment was analyzed using a binomial generalized linear mixed model with a logit link, using goal-attainment records (met/not met) from W2, W3, W4, W6, and W8, with results summarized over the study by goal and race. The model included week, goal, race, and their two- and three-way interactions. Pairwise comparisons were adjusted using the multivariate t method. *Significant Black vs. White difference for Goal 2, P < 0.05.

Panel C study food consumption was analyzed using a linear mixed-effects model as the total percentage of required intake days completed over the full 8-week intervention period for each food. The model included food, race, and their interaction. Pairwise comparisons were adjusted using the multivariate t method. Superscript letters in Panel C indicate adjusted pairwise differences among study foods; values sharing a letter are not significantly different, whereas values with different letters differ significantly at P < 0.05.

**Supplemental Table 3**. Acceptance and palatability scores for each study food by race over the whole 8-week intervention.

|  | **Acceptance*** | | **Palatability^¶^ *** | |
| --- | --- | --- | --- | --- |
| **Food** | **Black** | **White** | **Black** | **White** |
| **Almonds** | 8 [5.5, 9] | 7 [6, 8] | 69.7 ± 2.4 | 70.0 ± 1.4 |
| **Broccoli** | 7 [5.5, 8] | 6 [5, 7] | 66.0 ± 2.1 | 60.3 ± 1.6 |
| **Carrots** | 7 [6, 8] | 7 [6, 8] | 71.9 ± 1.6 | 67.7 ± 1.6 |
| **Clementines** | 8 [7, 9] | 7 [6, 8] | 82.5 ± 1.5 | 73.9 ± 1.8 |
| **Peppers** | 7 [6, 8] | 7 [5, 8] | 72.0 ± 2.0 | 68.6 ± 1.6 |
| **Prunes** | 7 [4, 8] | 6 [4, 7] | 62.8 ± 2.3 | 58.4 ± 2.1 |
| **Walnuts** | 6 [5, 8] | 6.5 [5, 8] | 61.5 ± 2.7 | 66.9 ± 1.6 |

Values are presented as median [IQR] for acceptance scores and non-transformed mean ± SE for palatability scores. ¶, analyses were conducted on transformed variable. *, Food x race interaction, P<0.05. Significant food x race interaction contrasts (P<0.05 after mvt adjustment for multiple comparisons) for acceptance scores were Clementines-Walnuts in Black vs White, OR=3.26, and Prunes-Walnuts in Black vs White, OR=4.20, while for palatability scores were Clementines-Walnuts in Black vs White, mean transformed estimate ± SE: 0.55 ± 0.18.

**Supplemental Table 4.** Self-reported dietary intake from the Diet History Questionnaire III for Black and White participants assigned to the PD and CD groups at Week 4 and Week 8, with adjustment for baseline intake and total energy intake

| Outcome | Week | PD |  | CD |  | BL-adjusted P-Values | | | | | | |
| --- | --- | --- | --- | --- | --- | --- | --- | --- | --- | --- | --- | --- |
|  |  | **Black** | **White** | **Black** | **White** | **G** | **R** | **W** | **G x R** | **W x G** | **W x R** | **W x G x R** |
| Diet quality indices subcomponent scores | | | | | | | | | | | | |
| AHEI total fruits score | W4 | 4.26 ± 0.3 | 4.23 ± 0.28 | 1.85 ± 0.32 | 1.39 ± 0.27 | **<0.001** | 0.694 | 0.875 | 0.264 | 0.700 | 0.451 | 0.774 |
|  | W8 | 4.17 ± 0.33 | 4.53 ± 0.28 | 1.71 ± 0.36 | 1.44 ± 0.27 |  |  |  |  |  |  |  |
| AHEI total nuts score | W4 | 9.3 ± 0.6 | 8 ± 0.6 | 4.86 ± 0.79 | 5.84 ± 0.59 | **<0.001** | 0.637 | 0.150 | 0.072 | 0.521 | 0.298 | 0.530 |
|  | W8 | 8.33 ± 0.66 | 8.35 ± 0.61 | 3.88 ± 0.64 | 5.24 ± 0.62 |  |  |  |  |  |  |  |
| AHEI total vegetables score ^a^ | W4 | 8.08 ± 0.39 | 7.03 ± 0.36 | 2.84 ± 0.39 | 3.38 ± 0.35 | <0.001 | 0.750 | 0.745 | **0.026** | 0.335 | 0.444 | 0.723 |
|  | W8 | 8.11 ± 0.39 | 7.5 ± 0.36 | 2.64 ± 0.39 | 3.34 ± 0.35 |  |  |  |  |  |  |  |
| DASH total fruit score | W4 | 3.77 ± 0.22 | 3.96 ± 0.19 | 2.38 ± 0.24 | 2.1 ± 0.2 | **<0.001** | 0.920 | 0.959 | 0.212 | 0.608 | 0.825 | 0.858 |
|  | W8 | 3.84 ± 0.21 | 4.04 ± 0.2 | 2.26 ± 0.25 | 2.09 ± 0.2 |  |  |  |  |  |  |  |
| DASH total nuts score | W4 | 3.6 ± 0.2 | 3.7 ± 0.2 | 2.4 ± 0.22 | 2.7 ± 0.2 | **<0.001** | 0.437 | 0.755 | 0.360 | 0.978 | 0.667 | 0.659 |
|  | W8 | 3.74 ± 0.25 | 3.63 ± 0.21 | 2.44 ± 0.25 | 2.75 ± 0.2 |  |  |  |  |  |  |  |
| DASH total vegetables score ^a^ | W4 | 4.28 ± 0.17 | 3.76 ± 0.17 | 1.87 ± 0.18 | 2.27 ± 0.17 | <0.001 | 0.889 | 0.488 | **0.009** | 0.850 | 0.659 | 0.748 |
|  | W8 | 4.29 ± 0.2 | 3.92 ± 0.17 | 1.91 ± 0.2 | 2.33 ± 0.17 |  |  |  |  |  |  |  |
| HEI-2015 total fruits score ^b^ | W4 | 4.37 ± 0.26 | 5.11 ± 0.24 | 3.52 ± 0.33 | 2.77 ± 0.24 | <0.001 | 0.956 | 0.713 | **0.001** | 0.824 | 0.940 | 0.857 |
|  | W8 | 4.38 ± 0.29 | 5.05 ± 0.24 | 3.41 ± 0.32 | 2.69 ± 0.25 |  |  |  |  |  |  |  |
| HEI-2015 total vegetables score | W4 | 4.89 ± 0.15 | 5 ± 0.15 | 3.34 ± 0.21 | 3.74 ± 0.16 | **<0.001** | 0.047 | 0.994 | 0.300 | 0.428 | 0.527 | 0.835 |
|  | W8 | 4.79 ± 0.17 | 4.97 ± 0.16 | 3.34 ± 0.21 | 3.88 ± 0.16 |  |  |  |  |  |  |  |
| MED total fruits score | W4 | 0.81 ± 0.07 | 0.94 ± 0.06 | 0.22 ± 0.08 | 0.1 ± 0.07 | **<0.001** | 0.995 | 0.833 | 0.104 | 0.783 | 0.949 | 0.346 |
|  | W8 | 0.85 ± 0.08 | 0.89 ± 0.07 | 0.2 ± 0.08 | 0.16 ± 0.07 |  |  |  |  |  |  |  |
| MED total legumes score | W4 | 0.37 ± 0.09 | 0.61 ± 0.09 | 0.5 ± 0.12 | 0.67 ± 0.09 | **<0.001** | 0.008 | 0.992 | 0.404 | 0.164 | 0.874 | 0.101 |
|  | W8 | 0.55 ± 0.1 | 0.6 ± 0.09 | 0.31 ± 0.1 | 0.69 ± 0.09 |  |  |  |  |  |  |  |
| MED total nuts score | W4 | 0.92 ± 0.07 | 0.74 ± 0.07 | 0.28 ± 0.09 | 0.15 ± 0.08 | **<0.001** | 0.244 | 0.567 | 0.494 | 0.469 | 0.159 | 0.798 |
|  | W8 | 0.85 ± 0.09 | 0.8 ± 0.07 | 0.26 ± 0.08 | 0.31 ± 0.08 |  |  |  |  |  |  |  |
| MED total vegetables score | W4 | 0.97 ± 0.06 | 0.85 ± 0.06 | 0.1 ± 0.06 | 0.11 ± 0.06 | **<0.001** | 0.861 | 0.369 | 0.065 | 0.169 | 0.223 | 0.450 |
|  | W8 | 0.94 ± 0.06 | 0.85 ± 0.06 | 0.11 ± 0.07 | 0.27 ± 0.07 |  |  |  |  |  |  |  |
| Macronutrients | | | | | | | | | | | | |
| Soluble fiber (g) | W4 | 6.57 ± 0.34 | 7.55 ± 0.34 | 4.75 ± 0.36 | 4.86 ± 0.34 | **<0.001** | 0.056 | 0.396 | 0.227 | 0.173 | 0.596 | 0.711 |
|  | W8 | 6.64 ± 0.36 | 7.68 ± 0.35 | 4.17 ± 0.4 | 4.62 ± 0.38 |  |  |  |  |  |  |  |
| Insoluble fiber (g) | W4 | 17.81 ± 0.8 | 16.73 ± 0.82 | 9.37 ± 0.82 | 8.35 ± 0.81 | **<0.001** | 0.202 | 0.729 | 0.984 | 0.399 | 0.902 | 0.968 |
|  | W8 | 18.29 ± 0.87 | 17.34 ± 0.88 | 9.1 ± 0.85 | 8.15 ± 0.82 |  |  |  |  |  |  |  |
| Total sugars (g) | W4 | 80.33 ± 5.08 | 86.7 ± 4.98 | 75.53 ± 7.02 | 67.16 ± 5.13 | **<0.001** | 0.377 | 0.447 | 0.241 | 0.162 | 0.100 | 0.586 |
|  | W8 | 78.59 ± 5.38 | 92.2 ± 5.24 | 61.72 ± 5.83 | 67.47 ± 5.97 |  |  |  |  |  |  |  |
| Micronutrients | | | | | | | | | | | | |
| Arginine (g) | W4 | 3.24 ± 0.2 | 3.23 ± 0.2 | 2.96 ± 0.23 | 3.33 ± 0.21 | **<0.001** | 0.813 | 0.904 | 0.575 | 0.384 | 0.299 | 0.429 |
|  | W8 | 3.4 ± 0.22 | 3.31 ± 0.2 | 3.09 ± 0.24 | 3.01 ± 0.24 |  |  |  |  |  |  |  |
| Beta-carotene (μg) ^a^ | W4 | 10618.56 ± 564.85 | 8074.47 ± 576.39 | 2084.38 ± 580.22 | 2197.95 ± 528.88 | <0.001 | 0.570 | 0.626 | **0.001** | 0.708 | 0.598 | 0.659 |
|  | W8 | 10674.28 ± 648.16 | 8931.93 ± 602.91 | 2144.17 ± 585.96 | 2674.92 ± 531.91 |  |  |  |  |  |  |  |
| Beta-cryptoxanthin (μg) | W4 | 486.76 ± 29.84 | 500.83 ± 29.14 | 69.36 ± 30.07 | 54.1 ± 27.76 | **<0.001** | 0.657 | 0.243 | 0.945 | 0.291 | 0.605 | 0.544 |
|  | W8 | 559.68 ± 48.27 | 525.91 ± 30.17 | 69.87 ± 30.59 | 58.73 ± 27.64 |  |  |  |  |  |  |  |
| Delta-tocopherol (mg) | W4 | 1.63 ± 0.18 | 1.48 ± 0.17 | 1.79 ± 0.19 | 1.69 ± 0.18 | **<0.001** | 0.222 | 0.630 | 0.253 | 0.270 | 0.498 | 0.081 |
|  | W8 | 1.44 ± 0.21 | 1.54 ± 0.18 | 2.23 ± 0.21 | 1.59 ± 0.18 |  |  |  |  |  |  |  |
| Cholesterol intake (mg) ^c^ | W4 | 140.77 ± 20.09 | 155.41 ± 20.56 | 173.36 ± 21.51 | 265.73 ± 22.38 | <0.001 | 0.121 | 0.722 | 0.403 | 0.808 | 0.037 | **0.043** |
|  | W8 | 145.92 ± 22.54 | 162.54 ± 20.92 | 219.57 ± 22.71 | 221.63 ± 22.35 |  |  |  |  |  |  |  |
| Conjugated linoleic acid (CLA) (g) | W4 | 0.05 ± 0.01 | 0.06 ± 0.01 | 0.08 ± 0.01 | 0.11 ± 0.01 | **<0.001** | 0.167 | 0.444 | 0.486 | 0.899 | 0.482 | 0.229 |
|  | W8 | 0.05 ± 0.01 | 0.06 ± 0.01 | 0.1 ± 0.01 | 0.1 ± 0.01 |  |  |  |  |  |  |  |
| Food (dietary) folate (μg) † | W4 | -2.17 ± 0.15 | -2.38 ± 0.15 | -3 ± 0.24 | -2.82 ± 0.15 | **<0.001** | 0.880 | 0.251 | 0.142 | 0.233 | 0.683 | 0.641 |
|  | W8 | -2.15 ± 0.17 | -2.36 ± 0.16 | -3.3 ± 0.22 | -2.96 ± 0.16 |  |  |  |  |  |  |  |
| Vitamin B6 (mg) † | W4 | 0.08 ± 0.13 | 0.16 ± 0.12 | -0.38 ± 0.15 | -0.05 ± 0.13 | **<0.001** | 0.071 | 0.104 | 0.285 | 0.073 | 0.784 | 0.974 |
|  | W8 | 0.07 ± 0.13 | 0.2 ± 0.13 | -0.66 ± 0.16 | -0.3 ± 0.15 |  |  |  |  |  |  |  |
| Lycopene (μg) | W4 | 2346.8 ± 430.1 | 2492.15 ± 404.01 | 3224.31 ± 444.75 | 3676.22 ± 400.87 | **<0.001** | 0.105 | 0.932 | 0.457 | 0.526 | 0.156 | 0.631 |
|  | W8 | 2252.06 ± 411.57 | 2824.08 ± 421.34 | 2640.92 ± 427.14 | 3946.18 ± 416.79 |  |  |  |  |  |  |  |
| Magnesium (mg) | W4 | 303.78 ± 16.69 | 314.71 ± 16.42 | 237.63 ± 17.71 | 248.64 ± 17.33 | **<0.001** | 0.636 | 0.535 | 0.606 | 0.215 | 0.729 | 0.442 |
|  | W8 | 321.81 ± 21.39 | 311.65 ± 16.61 | 213.43 ± 16.74 | 232.62 ± 18.08 |  |  |  |  |  |  |  |
| Manganese (mg) | W4 | 3.12 ± 0.2 | 2.99 ± 0.19 | 2.79 ± 0.23 | 2.3 ± 0.19 | **<0.001** | 0.143 | 0.155 | 0.561 | 0.243 | 0.792 | 0.470 |
|  | W8 | 3.16 ± 0.22 | 2.93 ± 0.19 | 2.4 ± 0.22 | 2.13 ± 0.2 |  |  |  |  |  |  |  |
| Phosphorus (mg) | W4 | 920.54 ± 53.26 | 995.79 ± 50.53 | 904 ± 58.95 | 1135.67 ± 54.49 | **<0.001** | **0.048** | 0.915 | 0.239 | 0.889 | 0.080 | 0.496 |
|  | W8 | 950.74 ± 61.13 | 967.55 ± 54.56 | 970.49 ± 62.31 | 1052.5 ± 67.14 |  |  |  |  |  |  |  |
| Potassium (mg) | W4 | 2567.65 ± 117.1 | 2647.68 ± 106.23 | 1812.88 ± 125.56 | 2067.85 ± 116.22 | **<0.001** | 0.220 | 0.597 | 0.551 | 0.268 | 0.676 | 0.678 |
|  | W8 | 2687.67 ± 114.9 | 2763.8 ± 116.05 | 1823.69 ± 159.35 | 1966.12 ± 133.49 |  |  |  |  |  |  |  |
| Retinol (μg) | W4 | 193.65 ± 25.65 | 247.2 ± 26.11 | 303.55 ± 37.58 | 384.07 ± 27.38 | **<0.001** | 0.124 | 0.329 | 0.744 | 0.782 | 0.081 | 0.700 |
|  | W8 | 204.56 ± 27.59 | 213.24 ± 26.03 | 317.19 ± 32.56 | 330.01 ± 27.61 |  |  |  |  |  |  |  |
| Total alpha-tocopherol (mg) | W4 | 14.4 ± 1.07 | 16.8 ± 1.14 | 8.99 ± 1.12 | 7.46 ± 1.02 | **<0.001** | 0.829 | 0.470 | 0.089 | 0.444 | 0.724 | 0.486 |
|  | W8 | 15.11 ± 1.26 | 16.25 ± 1.1 | 7.88 ± 1.08 | 6.71 ± 1.04 |  |  |  |  |  |  |  |
| Vitamin B12 ( μg) | W4 | 2.46 ± 0.34 | 2.93 ± 0.35 | 3.68 ± 0.43 | 4.91 ± 0.37 | **<0.001** | 0.055 | 0.059 | 0.251 | 0.483 | 0.380 | 0.863 |
|  | W8 | 2.37 ± 0.37 | 2.52 ± 0.34 | 3.4 ± 0.4 | 4.18 ± 0.38 |  |  |  |  |  |  |  |
| Vitamin C (mg) | W4 | 237.79 ± 12.63 | 235.5 ± 12.47 | 75.66 ± 12.99 | 60.51 ± 11.57 | **<0.001** | 0.716 | 0.711 | 0.720 | 0.223 | 0.482 | 0.705 |
|  | W8 | 241.59 ± 14.69 | 243.53 ± 12.24 | 57.19 ± 12.88 | 56.82 ± 11.42 |  |  |  |  |  |  |  |
| Vitamin D (μg)^d^ | W4 | 2.23 ± 0.46 | 2.73 ± 0.46 | 3.09 ± 0.49 | 4.83 ± 0.48 | **<0.001** | 0.178 | 0.404 | 0.267 | 0.861 | **0.014** | 0.479 |
|  | W8 | 2.45 ± 0.46 | 2.23 ± 0.46 | 3.53 ± 0.52 | 3.97 ± 0.49 |  |  |  |  |  |  |  |
| Food groups |  |  |  |  |  |  |  |  |  |  |  |  |
| Total protein foods (oz) | W4 | 4.68 ± 0.38 | 4.9 ± 0.4 | 3.95 ± 0.42 | 4.7 ± 0.42 | **<0.001** | 0.647 | 0.471 | 0.905 | 0.347 | 0.202 | 0.361 |
|  | W8 | 5.17 ± 0.46 | 5.24 ± 0.41 | 4.4 ± 0.44 | 4.1 ± 0.43 |  |  |  |  |  |  |  |
| Total fruit cups | W4 | 1.82 ± 0.14 | 1.77 ± 0.12 | 1.08 ± 0.18 | 0.68 ± 0.13 | **<0.001** | 0.369 | 0.400 | 0.103 | 0.385 | 0.177 | 0.912 |
|  | W8 | 1.7 ± 0.13 | 1.9 ± 0.13 | 0.82 ± 0.15 | 0.65 ± 0.12 |  |  |  |  |  |  |  |
| Total dairy cups | W4 | 0.69 ± 0.14 | 0.92 ± 0.14 | 1.17 ± 0.15 | 1.73 ± 0.16 | **<0.001** | 0.071 | 0.972 | 0.397 | 0.389 | 0.051 | 0.418 |
|  | W8 | 0.7 ± 0.14 | 0.77 ± 0.14 | 1.45 ± 0.17 | 1.6 ± 0.16 |  |  |  |  |  |  |  |
| Total grain (oz) | W4 | 2.94 ± 0.27 | 2.62 ± 0.26 | 4.37 ± 0.35 | 3.37 ± 0.28 | **<0.001** | 0.077 | 0.149 | 0.550 | 0.568 | 0.238 | 0.223 |
|  | W8 | 2.84 ± 0.28 | 2.51 ± 0.26 | 3.71 ± 0.35 | 3.45 ± 0.29 |  |  |  |  |  |  |  |
| Whole grain (oz) | W4 | 0.66 ± 0.12 | 0.53 ± 0.12 | 1.03 ± 0.17 | 0.57 ± 0.12 | **<0.001** | **0.040** | 0.050 | 0.441 | 0.230 | 0.447 | 0.282 |
|  | W8 | 0.63 ± 0.14 | 0.45 ± 0.12 | 0.63 ± 0.13 | 0.46 ± 0.12 |  |  |  |  |  |  |  |
| Refined grain (oz) | W4 | 2.2 ± 0.23 | 2.1 ± 0.23 | 3.22 ± 0.3 | 2.82 ± 0.24 | **<0.001** | 0.605 | 0.755 | 0.890 | 0.829 | 0.311 | 0.355 |
|  | W8 | 2.17 ± 0.25 | 2.09 ± 0.23 | 2.89 ± 0.26 | 3 ± 0.27 |  |  |  |  |  |  |  |

BL-adjusted emmeans ± SE are shown for W4 and W8 across Group x Race combinations. †Outcome was Johnson-transformed; transformed-scale emmeans shown. Repeated measures were analyzed using baseline-adjusted linear mixed-effects models with additional adjustment for total daily energy intake (fixed effects: G = group, W = week, R = race, GxR = group x race, GxW = group x week, WxR = week x race, WxGxR = week x group x race; random intercept: participant). PD = personalized whole foods group, CD = control group, W4 = week 4, W8 = week 8. Pairwise comparisons used multivariate-t (mvt) adjustment. All analyses followed a modified intention-to-treat (mITT) framework including all randomized participants with valid outcome data, with missing values addressed via multiple imputation.

Significant contrasts (P < 0.05): a, PD-CD Black>White; b, PD-CD Black<White; c, CD W4-W8: Black negative, White positive; d, W4-W8 Black negative, White positive

**Supplemental Table 5.** Physical activity, sleep, and appetite outcomes for Black and White participants in the PD and CD groups, with adjustment for baseline values.

| Outcome | Week | PD |  | CD |  | BL-adjusted P-Values | | | | | | |
| --- | --- | --- | --- | --- | --- | --- | --- | --- | --- | --- | --- | --- |
|  |  | **Black** | **White** | **Black** | **White** | **G** | **R** | **W** | **G x R** | **W x G** | **W x R** | **W x G x R** |
| Physical activity | | | | | | | | | | | | |
| Total activity time (min) | W4 | 33.78 ± 5.31 | 42.1 ± 5.04 | 38.75 ± 5.67 | 42.49 ± 5.11 | 0.657 | 0.435 | 0.262 | 0.975 | 0.794 | 0.416 | 0.506 |
|  | W8 | 35.87 ± 5.3 | 34.37 ± 4.92 | 34.71 ± 5.4 | 37.31 ± 5.03 |  |  |  |  |  |  |  |
| METs | W4 | 1.24 ± 0.04 | 1.28 ± 0.05 | 1.26 ± 0.04 | 1.25 ± 0.04 | 0.969 | 0.994 | 0.270 | 0.654 | 0.776 | 0.454 | 0.557 |
|  | W8 | 1.24 ± 0.04 | 1.22 ± 0.03 | 1.24 ± 0.04 | 1.22 ± 0.04 |  |  |  |  |  |  |  |
| Light activity time (min) | W4 | 101.18 ± 9.27 | 125.47 ± 8.88 | 116.45 ± 9.46 | 151 ± 9.43 | **0.011** | **0.010** | 0.506 | 0.466 | 0.945 | 0.152 | 0.924 |
|  | W8 | 114.65 ± 9.3 | 120.62 ± 8.57 | 128.05 ± 10 | 146.35 ± 11.45 |  |  |  |  |  |  |  |
| % time in light activity ^a, b^ | W4 | 14.47 ± 1.17 | 18.21 ± 1.14 | 17.39 ± 1.25 | 20.42 ± 1.14 | **0.003** | 0.083 | 0.859 | 0.437 | 0.904 | **0.029** | 0.615 |
|  | W8 | 15.97 ± 1.14 | 17.17 ± 1.03 | 19.42 ± 1.24 | 18.47 ± 1.3 |  |  |  |  |  |  |  |
| Sedentary time (min) | W4 | 544.08 ± 23.92 | 529.95 ± 24.88 | 567.07 ± 26.02 | 492.23 ± 22 | 0.267 | 0.166 | 0.408 | 0.833 | 0.460 | 0.249 | 0.113 |
|  | W8 | 578.8 ± 29.93 | 550.1 ± 23.08 | 522.6 ± 24.59 | 539.54 ± 23.92 |  |  |  |  |  |  |  |
| % time sedentary ^b, c^ | W4 | 81.16 ± 1.62 | 75.84 ± 1.54 | 79.34 ± 1.67 | 72.18 ± 1.59 | 0.071 | 0.004 | 0.986 | 0.483 | 0.694 | **0.015** | 0.979 |
|  | W8 | 78.45 ± 1.57 | 77.77 ± 1.45 | 77.32 ± 1.65 | 74.91 ± 1.65 |  |  |  |  |  |  |  |
| Moderate activity time (min) | W4 | 29.64 ± 5.09 | 39.07 ± 4.93 | 36.17 ± 4.95 | 41.95 ± 4.83 | 0.304 | 0.232 | 0.290 | 0.996 | 0.747 | 0.326 | 0.553 |
|  | W8 | 32.33 ± 4.74 | 31.73 ± 4.4 | 33.13 ± 4.83 | 36.26 ± 4.54 |  |  |  |  |  |  |  |
| % time in moderate activity | W4 | 4.29 ± 0.62 | 5.41 ± 0.66 | 5.32 ± 0.68 | 5.54 ± 0.62 | 0.330 | 0.774 | 0.304 | 0.511 | 0.808 | 0.195 | 0.770 |
|  | W8 | 4.6 ± 0.68 | 4.44 ± 0.59 | 5.2 ± 0.66 | 4.61 ± 0.61 |  |  |  |  |  |  |  |
| Vigorous activity time (min) | W4 | 3.52 ± 1.09 | 3 ± 0.93 | 1.66 ± 0.86 | 1.09 ± 0.8 | 0.102 | 0.942 | 0.286 | 0.886 | 0.199 | 0.414 | 0.850 |
|  | W8 | 1.62 ± 0.83 | 1.88 ± 0.81 | 1.19 ± 0.87 | 1.85 ± 0.82 |  |  |  |  |  |  |  |
| % time in vigorous activity | W4 | 0.55 ± 0.18 | 0.39 ± 0.12 | 0.23 ± 0.12 | 0.14 ± 0.11 | 0.104 | 0.821 | 0.278 | 0.718 | 0.119 | 0.279 | 0.986 |
|  | W8 | 0.22 ± 0.11 | 0.26 ± 0.11 | 0.17 ± 0.12 | 0.28 ± 0.12 |  |  |  |  |  |  |  |
| Sleep | | | | | | | | | | | | |
| Total Sleep Hours ^d^ | W4 | 6.71 ± 0.17 | 6.98 ± 0.17 | 6.76 ± 0.19 | 7.04 ± 0.17 | 0.335 | **<0.001** | 0.972 | 0.820 | 0.129 | **0.044** | 0.740 |
|  | W8 | 6.62 ± 0.2 | 7.48 ± 0.21 | 6.35 ± 0.21 | 7.06 ± 0.18 |  |  |  |  |  |  |  |
| Subjective appetite ratings ^^^ | | | | | | | | | | | | |
| Full day ratings | | | | | | | | | | | | |
| Hunger | W8 | 23392.2 ± 2448.86 | 25550.32 ± 2522.24 | 23225.98 ± 2455.03 | 23706.19 ± 2248.75 | 0.671 | 0.600 | — | 0.732 | — | — | — |
| Fullness | W8 | 36511.96 ± 2865.97 | 40533.85 ± 2642.4 | 38860.31 ± 3051.02 | 44019.18 ± 2577.77 | 0.290 | 0.109 | — | 0.835 | — | — | — |
| Desire to eat | W8 | 23216.83 ± 2342.6 | 24891.92± 2367.51 | 22537.22 ± 2297.51 | 24104.31 ± 2146.74 | 0.750 | 0.472 | — | 0.981 | — | — | — |
| Prospective consumption | W8 | 22571.7 ± 2109.36 | 27144.64 ± 1983.97 | 25752.42 ± 2095.92 | 27575.26 ± 2002.63 | 0.380 | 0.118 | — | 0.507 | — | — | — |
| Morning ratings | | | | | | | | | | | | |
| Hunger | W8 | 7825.67 ± 945.37 | 7914.01 ± 968.72 | 7533.39 ± 1069.66 | 6132.28 ± 931.95 | 0.294 | 0.519 | — | 0.436 | — | — | — |
| Fullness | W8 | 8830.63 ± 1115.12 | 10682.71 ± 1054.56 | 8996.09 ± 1156.72 | 12039.41 ± 1028.72 | 0.488 | **0.025** | — | 0.587 | — | — | — |
| Desire to eat | W8 | 7889.65 ± 994.19 | 7623.1 ± 918.26 | 6586.99 ± 1036.69 | 6699.59 ± 892.65 | 0.249 | 0.937 | — | 0.841 | — | — | — |
| Prospective consumption | W8 | 8024.79 ± 927.25 | 8467.8 ± 854.49 | 7441.72 ± 937.79 | 7769.92 ± 831.01 | 0.482 | 0.661 | — | 0.950 | — | — | — |
| Afternoon ratings | | | | | | | | | | | | |
| Hunger | W8 | 8204.32 ± 1077.6 | 8862.27 ± 1032.82 | 8229.37 ± 1090.64 | 8559.28 ± 1021.38 | 0.894 | 0.646 | — | 0.880 | — | — | — |
| Fullness | W8 | 13998.76 ± 1285.03 | 15287.54 ± 1191.67 | 12906.62 ± 1285.82 | 16251.62 ± 1152.6 | 0.959 | 0.065 | — | 0.396 | — | — | — |
| Desire to eat | W8 | 8508.33 ± 1069.73 | 7930.21 ± 940.05 | 8323.48 ± 1072.66 | 8388.05 ± 904.69 | 0.892 | 0.800 | — | 0.748 | — | — | — |
| Prospective consumption | W8 | 8837.43 ± 1148.39 | 9154 ± 952.52 | 9686.25 ± 1031.99 | 9923.83 ± 945.5 | 0.427 | 0.782 | — | 0.970 | — | — | — |
| Evening ratings | | | | | | | | | | | | |
| Hunger | W8 | 7279.62 ± 1068.58 | 8629.59 ± 1052.57 | 7396.34 ± 1124.47 | 9245.22 ± 1067.71 | 0.730 | 0.133 | — | 0.817 | — | — | — |
| Fullness | W8 | 14186.44 ± 1200.42 | 15002.37 ± 1171.01 | 17049.21 ± 1294.99 | 16037.56 ± 1173.93 | 0.109 | 0.935 | — | 0.458 | — | — | — |
| Desire to eat | W8 | 6487.74 ± 1092.47 | 8919.26 ± 1086.77 | 7833.35 ± 1139.69 | 9282.98 ± 1133.04 | 0.434 | 0.097 | — | 0.651 | — | — | — |
| Prospective consumption | W8 | 6346.85 ± 889.4 | 9677.34 ± 844.82 | 8149.16 ± 944.46 | 9466.74 ± 826.94 | 0.373 | **0.008** | — | 0.265 | — | — | — |

BL-adjusted emmeans ± SE are shown for W4 and W8 across Group x Race combinations. ^Outcome was measured at baseline and W8 only. Outcomes assessed at BL, W4, and W8 (physical activity, sleep) were analyzed using baseline-adjusted linear mixed-effects models (fixed effects: G = group, W = week, R = race, G x R = group x race, G x W = group x week, W x R = week x race, W x G x R = week x group x race; random intercept: participant). Physical activity outcomes were additionally adjusted for total accelerometer wear time. Sleep midpoint represents average of W4 and W5 measurements, with baseline as W1 average. Outcomes assessed at BL and W8 (appetite ratings) were analyzed using baseline-adjusted linear models (fixed effects: G, R, G x R; baseline value as covariate). PD = personalized whole foods group, CD = control group, W4 = week 4, W8 = week 8. Pairwise comparisons used multivariate-t (mvt) adjustment. All analyses followed a modified intention-to-treat (mITT) framework including all randomized participants with valid outcome data, with missing values addressed via multiple imputation.

Significant contrasts (P<0.05): a, W4-W8: Black negative, White positive; b, W4: Black vs White; c, W4-W8: Black positive, White negative; d, W8: Black < White

**Supplemental Table 6.** Glucoregulation-associated serum markers for Black and White participants in the PD and CD groups, with baseline-adjusted fasting measures at Week 4 and Week 8 and baseline-adjusted OGTT-derived measures at Week 8.

| Outcome | Week | PD | | CD | | BL-adjusted P-Values | | | | | | |
| --- | --- | --- | --- | --- | --- | --- | --- | --- | --- | --- | --- | --- |
|  |  | **Black** | **White** | **Black** | **White** | **G** | **R** | **W** | **G x R** | **W x G** | **W x R** | **W x G x R** |
| Fasting measures | | | | | | | | | | | | |
| ‡ Fasting insulin (µU/mL) ^a, b^ | W4 | 18.95 ± 1.24 | 17.22 ± 1.23 | 18.96 ± 1.35 | 17.48 ± 1.2 | 0.859 | 0.630 | 0.002 | 0.165 | 0.970 | 0.201 | **0.050** |
|  | W8 | 13.98 ± 1.27 ^c^ | 17.31 ± 1.24 | 17.09 ± 1.41 | 14.59 ± 1.25 |  |  |  |  |  |  |  |
| Fasting blood glucose (mg/dL) | W4 | 81.79 ± 1.7 | 82.88 ± 1.64 | 81.79 ± 1.75 | 80.41 ± 1.66 | 0.960 | 0.844 | **<0.001** | 0.197 | 0.096 | 0.584 | 0.373 |
|  | W8 | 76.16 ± 1.74 | 79.59 ± 1.65 | 80.25 ± 1.75 | 78.29 ± 1.72 |  |  |  |  |  |  |  |
| Fasting NEFA (mmol/L) | W4 | 0.41 ± 0.04 | 0.36 ± 0.03 | 0.39 ± 0.04 | 0.37 ± 0.03 | 0.661 | 0.902 | 0.324 | 0.953 | 0.474 | 0.168 | 0.503 |
|  | W8 | 0.33 ± 0.04 | 0.37 ± 0.03 | 0.37 ± 0.04 | 0.38 ± 0.03 |  |  |  |  |  |  |  |
| HOMA-IR | W4 | 3.89 ± 0.27 | 3.66 ± 0.26 | 3.82 ± 0.29 | 3.54 ± 0.27 | 0.902 | 0.876 | **<0.001** | 0.131 | 0.461 | 0.208 | 0.076 |
|  | W8 | 2.65 ± 0.26 | 3.44 ± 0.27 | 3.41 ± 0.28 | 2.97 ± 0.27 |  |  |  |  |  |  |  |
| HOMA-β† | W4 | -0.13 ± 0.17 | -0.24 ± 0.17 | 0.16 ± 0.18 | 0.06 ± 0.16 | **0.030** | 0.829 | 0.657 | 0.605 | 0.897 | 0.471 | 0.407 |
|  | W8 | -0.25 ± 0.18 | -0.07 ± 0.17 | 0.22 ± 0.18 | 0.1 ± 0.19 |  |  |  |  |  |  |  |
| OGTT measures ^^^ | | | | | | | | | | | | |
| Matsuda Index (MI) | W8 | 4.5 ± 0.67 | 3.3 ± 0.62 | 3.4 ± 0.7 | 3.57 ± 0.69 | 0.494 | 0.531 | — | 0.310 | — | — | — |
| Glucose AUCs ^^^ | | | | | | | | | | | | |
| 0–15 min ^b, d^ | W8 | 1344.81 ± 27.68 | 1500.49 ± 27.12 | 1423.71 ± 29.16 | 1427.45 ± 26.84 | 0.918 | **0.005** | — | 0.008 | — | — | — |
| 0–30 min ^b, d^ | W8 | 3074.08 ± 73.6 | 3464.24 ± 68.05 | 3219.43 ± 71.1 | 3302.78 ± 70.25 | 0.912 | **0.001** | — | **0.036** | — | — | — |
| 0–60 min | W8 | 6849.75 ± 161.16 | 7440.12 ± 151.81 | 7030.91 ± 168.4 | 7217.15 ± 155.26 | 0.892 | **0.017** | — | 0.201 | — | — | — |
| 0–120 min | W8 | 13630.45 ± 335.27 | 14700.25 ± 337.15 | 13898.88 ± 377.14 | 14074.16 ± 372.39 | 0.605 | 0.106 | — | 0.210 | — | — | — |
| Insulin AUCs ^^^ | | | | | | | | | | | | |
| 0–15 min | W8 | 707.76 ± 76.72 | 887.14 ± 72.04 | 866.72 ± 83.59 | 868.67 ± 76.17 | 0.317 | 0.329 | — | 0.244 | — | — | — |
| 0–30 min | W8 | 2346.41 ± 212.12 | 2926.69 ± 205.55 | 2544.5 ± 231.16 | 2730.13 ± 202.9 | 0.997 | 0.132 | — | 0.348 | — | — | — |
| 0–60 min | W8 | 6408.38 ± 472.19 | 7129.5 ± 447.61 | 6374.7 ± 499.98 | 6656.4 ± 449.51 | 0.561 | 0.353 | — | 0.641 | — | — | — |
| 0–120 min | W8 | 12990.2 ± 1044.77 | 13291.3 ± 895.16 | 13014.2 ± 998.63 | 12462.85 ± 913.55 | 0.645 | 0.915 | — | 0.645 | — | — | — |
| NEFA AUCs ^^^ | | | | | | | | | | | | |
| 0–15 min† | W8 | -0.03 ± 0.2 | -0.02 ± 0.19 | 0.04 ± 0.2 | -0.1 ± 0.18 | **0.969** | 0.748 | — | 0.682 | — | — | — |
| 0–30 min† | W8 | -1.25 ± 0.19 | -1.36 ± 0.16 | -1.26 ± 0.17 | -1.46 ± 0.16 | 0.732 | 0.409 | — | 0.793 | — | — | — |
| 0–60 min† | W8 | -0.81 ± 0.18 | -1.01 ± 0.18 | -0.86 ± 0.19 | -1.11 ± 0.17 | 0.673 | 0.267 | — | 0.887 | — | — | — |
| 0–120 min† | W8 | 0.11 ± 0.2 | -0.1 ± 0.19 | 0.05 ± 0.19 | -0.23 ± 0.18 | 0.583 | 0.263 | — | 0.851 | — | — | — |

BL-adjusted emmeans ± SE are shown for W4 and W8 across Group x Race combinations. † Outcome was Johnson-transformed; transformed-scale emmeans shown. ‡ For this outcome, the Week x Group x Race interaction reached significance only in the sensitivity analysis adjusting for baseline BMI and food security score (P<0.05); emmeans ± SE and P-values shown are from that sensitivity analysis. Fasting measures (assessed at BL, W4, and W8) were analyzed using baseline-adjusted linear mixed-effects models (fixed effects: G = group, W = week, R = race, G x R = group x race, G x W = group x week, W x R = week x race, W x G x R = week x group x race; random intercept: participant). OGTT-derived measures (assessed at BL and W8) were analyzed using baseline-adjusted linear models (fixed effects: G, R, G x R; baseline value as covariate). ^Outcome was measured at baseline and W8. PD = personalized whole foods group, CD = control group, W4 = week 4, W8 = week 8. Pairwise comparisons used multivariate-t (mvt) adjustment. All analyses followed a modified intention-to-treat (mITT) framework including all randomized participants with valid outcome data, with missing values addressed via multiple imputation.

Significant contrasts: a, PD W4-W8 Black>White; b, W8 PD-CD: Black negative, White positive; c, PD Black: W8<W4; d, PD: Black<White.

**Supplemental Table 7.** Cognitive function outcomes for Black and White participants in the PD and CD groups at Week 8, with adjustment for baseline values.

| Outcome | Week | PD |  | CD |  | BL-adjusted P-values | | |
| --- | --- | --- | --- | --- | --- | --- | --- | --- |
|  |  | Black | White | Black | White | G | R | G x R |
| Concentration Performance (CP) | W8 | 175 ± 4.01 | 169.84 ± 3.67 | 160.74 ± 3.73 | 166.38 ± 3.67 | **0.019** | 0.951 | 0.158 |
| Total items processed (TN) ^a^ | W8 | 404.26 ± 9.42 ^b^ | 389.82 ± 8.22 | 361.07 ± 8.42 | 388.46 ± 8.21 | 0.010 | 0.458 | **0.016** |
| TN minus errors (TNE) ^a^ | W8 | 397.04 ± 8.02 ^b^ | 383.45 ± 7.87 | 359.62 ± 8.23 | 378.84 ± 7.68 | 0.010 | 0.728 | **0.041** |
| Memory (10-word recall total) | W8 | 7.13 ± 0.3 | 6.72 ± 0.25 | 6.97 ± 0.27 | 6.45 ± 0.25 | 0.429 | 0.090 | 0.831 |

BL-adjusted emmeans ± SE are shown for W8 across Group x Race combinations. Outcomes were analyzed using baseline-adjusted linear models (fixed effects: G = group, R = race, G x R = group x race; baseline value as covariate). PD = personalized whole foods group, CD = control group, W8 = week 8. Pairwise comparisons used multivariate-t (mvt) adjustment. All analyses followed a modified intention-to-treat (mITT) framework including all randomized participants with valid outcome data, with missing values addressed via multiple imputation.

Significant contrasts: a, PD-CD: Black > White; b, W8 Black: PD > CD.
